# Supporting women, girls and people who menstruate to participate in physical activity – Rapid evidence summary

**DOI:** 10.1101/2024.09.02.24311982

**Authors:** Judit Csontos, Deborah Edwards, Elizabeth Gillen, Juliet Hounsome, Meg Kiseleva, Mala Mann, Amrita Sidhu, Steven Macey, Ruth Lewis, Adrian Edwards, Alison Cooper

## Abstract

Insufficient physical activity is a leading risk factor for non-communicable diseases and has a negative effect on mental health and quality of life. Women, girls and people who menstruate living in Wales are less likely to engage in regular physical activity than boys and men. The aim of this rapid evidence summary is to identify research focusing on physical activity participation (including exercise and sport) of women, girls and people who menstruate in relation to the menstrual cycle, to inform the Welsh Government Period Proud Action Plan.

**Results:** 42 reports were identified, including overviews of reviews, systematic reviews, a scoping review, organisational reports, and primary studies. The secondary research evidence was published between 2008 and 2024 with the most recent searches being conducted in September 2023. The primary studies were published between 2020 and 2022. The organisational reports were published between 2018 and 2024.

**Research Implications and Evidence Gaps:** There is a need for interventions that could support physical activity participation (including exercise or sport) of women, girls and people who menstruate in relation to the menstrual cycle. These interventions need to consider and address barriers that women, girls and people who menstruate face in relation to their menstrual cycle, and robust evaluations are required to determine effectiveness.

**Funding statement:** The Cardiff Evidence Synthesis Collaborative were funded for this work by the Health and Care Research Wales Evidence Centre, itself funded by Health and Care Research Wales on behalf of Welsh Government.

## 1. CONTEXT / BACKGROUND

Insufficient physical activity (PA) is a leading risk factor for non-communicable diseases and has a negative effect on mental health and quality of life (Guthold et al. 2018). Women living in Wales are less likely to engage in regular PA (150 min/week moderate-intensity, 75 min/week vigorous-intensity PA, or any combination of these) than men, in line with global trends (Sport Wales 2021, Guthold et al. 2018). Adolescent girls are less active than boys, with 84% reporting reduced interest in sport and activities following menarche and 23% feel embarrassed to participate in PA during their periods (Nuffield Health 2023). Associated menstrual symptoms can also be a barrier to PA, such as pain and discomfort (House of Commons 2024). Trans and non-binary populations can experience additional challenges related to menstruation, including the use of menstrual products and men’s toilets (Frank 2020), which may also influence their PA participation. However, current NHS guidance does not include any advice on menstrual health during PA for women, girls and people who menstruate (NHS 2024). The Welsh Government is committed to promoting equity and inclusion in health and social care, as outlined in The Quality Statement for women and girls’ health (Welsh Government 2022). Therefore, the aim of this rapid evidence summary is to identify research focusing on the PA participation (including exercise and sport) of women, girls and people who menstruate in relation to the menstrual cycle, to inform the Welsh Government Period Proud Action Plan (Welsh Government 2023).

## 2. RESEARCH QUESTION(S)

The original research question suggested by the stakeholder group was: What is the impact of periods on women, girls and people who menstruate in relation to participation in sport/exercise and what innovations/interventions could support/encourage participation?

Due to the broad scope of this research question, the Population, concept, context (PCC) pneumonic was used to develop the rapid evidence summary research question and search the evidence base (Peters et al. 2020).

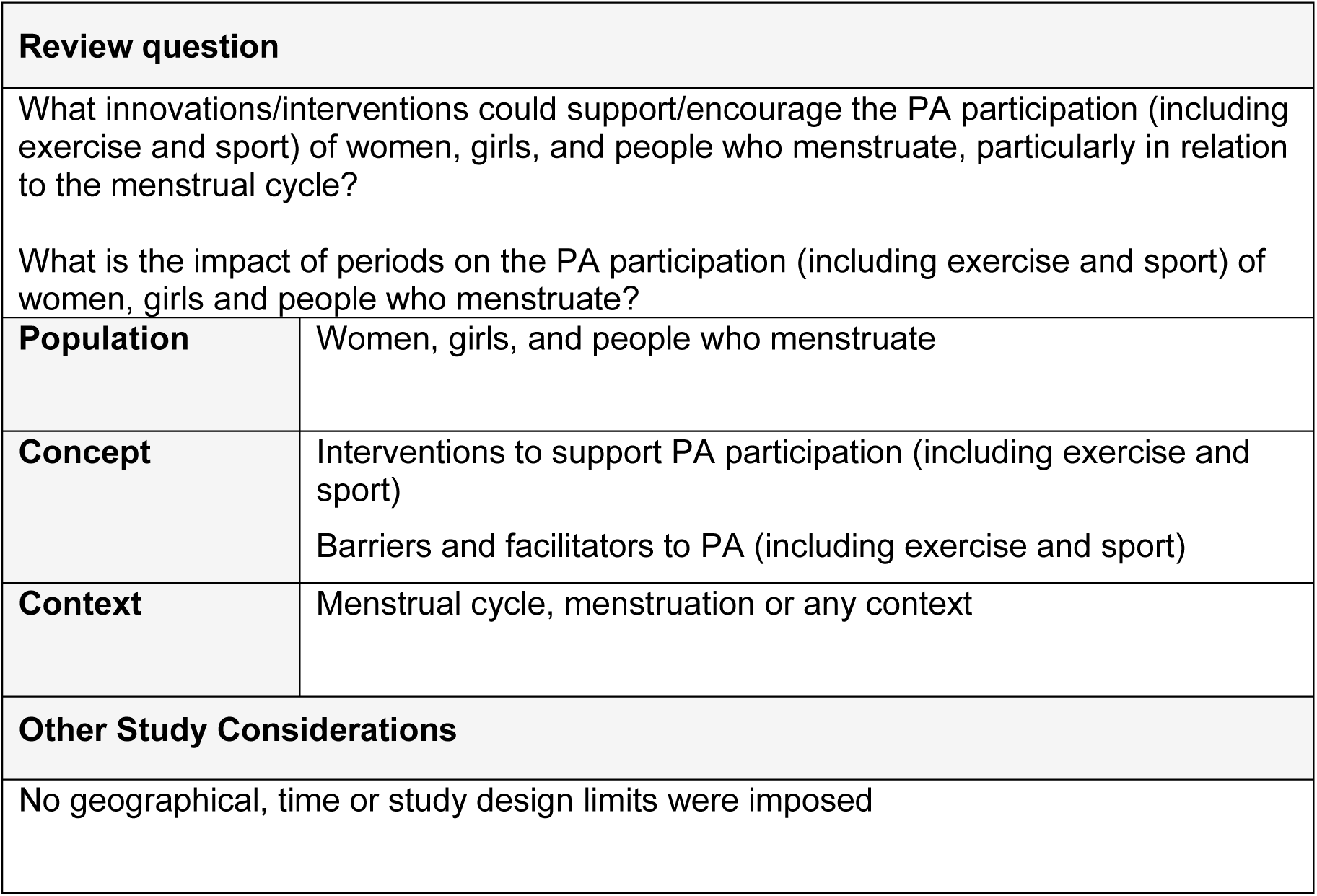

## 3. SUMMARY OF THE EVIDENCE BASE

### 3.1. Type and amount of evidence available

Overviews of reviews (n=2), systematic reviews (n=25), a scoping review (n=1), organisational reports (n=7), and primary research studies (n=7) were included in this preliminary literature review to provide a brief overview of the available evidence. As limited evidence was found on the topic of interventions to support PA participation in relation to menstruation (primary studies n=2, organisational report n=1), additional searches for evidence in the following topics were conducted: barriers to PA in relation to the menstrual cycle; wider barriers to PA participation of women, girls, and people who menstruate; interventions (unrelated to managing the menstrual cycle) to support PA participation; the effects of PA on dysmenorrhea and premenstrual syndrome (PMS); and the menstrual cycle and exercise performance. Throughout this rapid evidence summary, multiple terms relating to different types of PA, exercise or sport is mentioned. While multiple definitions may exist for these concepts (WHO 2020), below the evidence is summarised by using the terms the original authors of the included evidence adopted.

#### 3.1.1. Barriers to physical activity in relation to the menstrual cycle

Two systematic reviews were identified with a focus on:

- Factors in a sports environment which could facilitate or hinder cyclically menstruating athletes’ healthy sports participation in relation to their menstrual cycles (Srinivasa Gopalan et al. 2024a).
- The impact of symptoms, experiences and perceptions of the menstrual cycle on recreational PA behaviours in the general population (Srinivasa Gopalan et al. 2024b).

Six organisational reports were identified from the grey literature, and these focused on:

- The health barriers to sport in two groups in the UK: school-aged girls and women experiencing menopausal and perimenopausal symptoms (House of Commons 2024).
- Girls’ experiences of menstruation in the UK and explored the impact of menstruation on sport and exercise (Plan International UK 2018).
- Female participation in sport and PA in Scotland and discussed barriers faced by adolescent girls and women of all ages including menstruation (The Scottish Parliament: Health, Social Care and Sport Committee 2023).
- Supporting women aged 41-60 in the UK to be active during midlife and perimenopause/menopause, while exploring barriers to PA participation (Women in Sport 2021).
- Tackling adolescent girls’ disengagement from sport and exercise in the UK while also considering the barrier of periods (Women in Sport 2022).
- School-aged girls’ participation in sport and PAs in the UK, while it examined barriers to PA at school and physical education (PE) in relation to the menstrual cycle. Girls’ ideas were also investigated regarding what schools could do to help them in PE during their menstruation (Youth Sport Trust 2023).

Five primary studies were identified:

- A UK qualitative study that explored adolescent girls’ views on how menstruation influences their PA (Harvey et al. 2020).
- A cross-sectional study that investigated the relationship between menstruation and leisure time PA in female university students in South Korea (Kim et al. 2022).
- A mixed-methods study that aimed to quantify events of the menstrual cycle and self-reported PA avoidance in women in the UK; and to understand women’s lived experiences of PA throughout the menstrual cycle (Kolic et al. 2021).
- A mixed-methods study that assessed the impact of the menstrual cycle on football performance and exercise ability, and identified barriers in relation to menstruation in amateur female footballers in the UK (Pinel et al. 2022).
- A mixed-methods study that examined the effect of menstruation and PMS on habitual participation in adventurous activities (mountaineering, wild camping, running, wild swimming, climbing, etc.) in the UK (Prince & Annison 2023).

#### 3.1.2. Interventions to increase physical activity participation in relation to the menstrual cycle

Two primary studies were identified:

- A quasi-experimental study that examined the effects of social-media-based support on PMS and PA among female university students in South Korea (Nam & Cha 2020).
- A qualitative study that reported the results of a participatory project consisting of two workshops where adolescent participants in Sweden designed menstrual technologies (Sondergaard et al. 2021).

One organisational report was identified, which focused on:

- The impact of the “Big Sister: By girls for Girls” project which aimed to empower and support adolescent girls in the UK to enjoy sport, exercise and PA during puberty. A part of the project was “Hey Girls” which provided free sanitary products for girls in areas of deprivation (Women in Sport 2023).

#### 3.1.3. Wider barriers to physical activity participation of women, girls, and people who menstruate

One overview of reviews was identified, and it focused on:

- Summarising the barriers and facilitators of PA participation in adolescent girls from existing systematic reviews and identifying key areas for fostering gender-responsive action and policy implications (Duffey et al. 2021).

Six systematic reviews were identified, and these focused on:

- Synthesising evidence from qualitative studies relating to adolescent girls’ perceptions of PA participation (Corr et al. 2018).
- Synthesising current literature on barriers to exercise in postpartum women (Edie et al. 2021).
- Investigating pregnant women’s attitudes and perceived barriers and enablers to PA, including the perspectives of women diagnosed with gestational diabetes mellitus (Harrison et al. 2018).
- Examining factors that influence adolescent girls’ participation in sports and categorising these factors into constructs of the planned behaviour theory (Hopkins et al. 2022).
- Synthesising self-identified barriers and facilitators to young adult women’s participation in PA and offering suggestions for future studies in this population (Peng et al. 2023).
- Investigating influencing factors and adaptation approaches of PA interventions among midlife immigrant women (Zou et al. 2021).

#### 3.1.4. Interventions (unrelated to managing the menstrual cycle) to support physical activity participation of women, girls, and people who menstruate

Eight systematic reviews were identified, and these focused on:

- Assessing the impact of PA interventions on adolescent girls and young adults participation in team sport and identifying potential strategies for increasing participation (Allison et al. 2017).
- Evaluating the effectiveness, acceptability, and active behavioural change techniques of mobile PA technologies among midlife women undergoing menopause (AlSwayied et al. 2022).
- Examining the effectiveness of interventions to increase PA among pre-adolescent girls aged 5–11 years (Biddle et al. 2014).
- Describing the available evidence from PA interventions that targeted young and adolescent girls and determining their effectiveness and key characteristics of success (Camacho-Minano et al. 2011).
- Investigating the relationship between social support and PA in adolescent girls aged 10-19 years, and exploring how different types and providers of social support might influence this relationship (Laird et al. 2016).
- Examining interventions targeting the intersection of body image and movement experiences among girls and women (Matheson et al. 2023).
- Examining the effectiveness of interventions for increasing levels of PA and core physical skills in adolescent girls aged 11-18 years (NICE 2008).
- Investigated the implementation and outcomes of telehealth exercise interventions in the postpartum population (Turner et al. 2023).

#### 3.1.5. The effects of physical activity on dysmenorrhea

Three systematic reviews were identified, and these focused on:

- Investigating the effectiveness and safety of exercise for primary dysmenorrhoea (Armour et al. 2019a).
- Analysing the evidence for participant led self-care and lifestyle techniques in primary dysmenorrhea (Armour et al. 2019b).
- Evaluating the effectiveness of PA for the treatment of primary dysmenorrhea (Matthewman et al. 2018).

#### 3.1.6. The effects of exercise on premenstrual syndrome

Two systematic reviews were identified, and these focused on:

- Examining the evidence for the effectiveness of exercise as a treatment for PMS (Pearce et al. 2020).
- Investigating the effect of exercise on premenstrual symptoms (Saglam & Orsal 2020).

#### 3.1.7. The menstrual cycle and exercise performance

One overview of reviews was identified, and it focused on:

- Examining and critically evaluating the influence of menstrual cycle phase on acute performance and chronic adaptations to resistance exercise training (Colenso-Semple et al. 2023).

One scoping review were identified, and it focused on:

- Providing a summary of the biological mechanisms underlying the menstrual cycle’s impact on various performance determining anatomical and physiological parameters, and to identify various proposed concepts and theories that may explain performance changes following hormonal fluctuations (Bernstein & Behringer 2023).

Four systematic reviews were identified, and these focused on:

- Determining the effects of the menstrual cycle on exercise performance and provide evidence-based, practical recommendations to eumenorrheic women (McNulty et al. 2020).
- Establishing a consensus across studies to enable evidence-based recommendations for training individualisation depending on menstrual cycle phases (Meignié et al. 2021).
- Examining the effects of the different menstrual cycle phases on isometric, isokinetic, and dynamic maximum strength in female athletes (Niering et al. 2024).
- The impact of the menstrual cycle phase on perceived exertion during aerobic exercise in eumenorrheic women (Prado et al. 2023).

### 3.2. Key Findings

This section summarises the key findings of the identified reviews and studies. Additionally, more information about the included evidence can be found in Tables 2, 3 and 4. Table 2 summarises the identified secondary research evidence (systematic and scoping reviews, and overviews of reviews). Table 3 contains details of primary research studies, while Table 4 presents information extracted from organisational reports.

#### 3.2.1. Barriers to exercising in relation to the menstrual cycle

- A systematic review revealed that **cyclically menstruating athletes (amateur to elite level)** and their coaches experienced **barriers to sport participation**, such as menstruation taboo, lack of knowledge and awareness, and lack of communication among stakeholders. Facilitators included female coaches, positive experiences of communicating about the menstrual cycle, and trust (Srinivasa Gopalan et al. 2024a).
- A systematic review which focused on the **general population** found **that adult women and adolescent girls** experience **barriers to recreational PA participation**, such as sociocultural taboo against menstruation, lack of knowledge and resources related to the menstrual cycle. An individualised approach to recreational PA promotion is needed to account for variability in menstrual symptoms and experiences (Srinivasa Gopalan et al. 2024b).
- A House of Commons (2024) report suggested that **barriers** to **school-aged girls’** sport and exercise participation were partly driven by hormonal differences. The **barriers** included: difference between girls’ and boys’ puberty experiences; lack of timely school education on managing the menstrual cycle; insufficient training for teachers on how to educate on the menstrual cycle; PE teachers’ inadequate understanding on girls’ experiences of puberty. One highlighted recommendation to increase participation in PE was that schools should allow a wide choice of kit for girls to improve comfort. Identified **barriers** for **women’s (aged 41-60)** participation included perimenopausal and menopausal symptoms.
- An organisational report, which focused on **girl’s experience of menstruation** in the UK, identified the universal issues of stigma, taboo, access to menstrual products, and pain management. The main **barrier to sport and exercise participation** was concern over leaking (Plan International UK 2018).
- The Scottish Parliament: Health, Social Care and Sport Committee (2023) reported that there was a persistent gender gap in the rates of participation in sport and PA. **Barriers** faced by **adolescent girls** included a lack understanding of the impact that menstruation can have on their participation in PA. Further barriers included access and appropriateness of toilet/changing facilities and sports kit. The Scottish Government’s Women’s Health Plan encourages education about menstrual health as part of the Scottish curriculum, although evaluation of this plan has not been undertaken. Additionally, new statutory guidance on school uniforms could address issues with sport kit. **Barriers** faced by **women of all ages** included period pain, leakage, and lack of adequate toilet facilities in sports environments.
- An organisational report from Women in Sport (2021) found that approximately one third of **women aged 41-60** did not meet PA guidelines. Based on a survey, while 30% of women reported lower activity levels since menopause, 71% of women wanted to be more active. Ninety percent of respondents would be more open to the idea if PA was prescribed by a GP or healthcare professional. Reported **barriers** included limited perception of what PA could entail, fear of standing out and judgement. Suggested interventions included creating support networks, expanding their frame of reference on PA, and engaging health professionals and employers to help encourage women to take up PA in midlife.
- An organisational report of a large survey including over 4,000 **adolescent boys and girls** identified a gender gap regarding disengagement from sport and exercise, with 43% of girls saying that they used to be sporty but no longer are compared to 24% of boys (Women in Sport 2022). **Barriers faced by girls** included self-belief, capability, body image and periods. Around 7 in 10 avoided being active when on their period, with **barriers** including pain (73%), fear of leakage (62%), tiredness (52%) and self-consciousness (45%).
- An organisational report from Youth Sport Trust (2023) highlighted that 38% of **secondary school aged girls** reported their **periods as a barrier to participating in PA** in school, and 21% reported it out of school. Overall, periods were the most reported **barrier**. Only 51% of girls reported that they were comfortable to take part in PE whilst on their period. Almost half of the girls surveyed (48%) reported that they were not at all comfortable with discussing their period with their teacher, with 8% reporting that they were comfortable to do so. Girls’ main concerns about participating in PE or school sport whilst on their period were: pain and discomfort (68%), leaking (60%), and low mood (57%). When asked about how school could help them take part in PE whilst on their period, 40% requested teachers to have more empathy and understanding. Over half of the responding girls (56%) wanted more options in PE kit.
- A qualitative study conducted by Harvey et al. (2020) identified **barriers to adolescent girls’ PA participation during menstruation**, such as feeling unprepared due to insufficient information about menstruation received too late (following menarche); fear of leaking; feeling exposed and uncomfortable; clothing; disliking tampons; and menstrual symptoms. Facilitators included social influences, shared responsibility with peers in sports, and being active helped relieve premenstrual symptoms. Participants reported diverse strategies to overcome barriers including wearing dark clothing, carrying spare underwear, choosing lower intensity activities, among others.
- A cross-sectional survey of **female university students in South Korea** found that those with regular menstruation patterns had higher health consciousness, greater level of PA and intention to participate in **leisure time PA** than those with irregular periods (Kim et al. 2022).
- A mixed methods study of **women’s experiences** revealed that those who avoided PA when menstruating reported heavier periods, higher levels of fatigue and pain compared to non-avoiders (Kolic et al. 2021). **Reasons for PA avoidance** or modification included menstrual symptoms, personal thoughts, and concerns about other people’s views on menstruation.
- A mixed methods study, which focused on **amateur female footballers,** revealed that 73% of participants felt that menstruation presented **one or more barriers** to football participation. Confidence and aerobic capacity were identified to be the most negatively impacted during pre-menstrual and menstrual stages (Pinel et al. 2022).
- A mixed methods study of **women attending adventurous activities (mountaineering, wild camping, running, wild swimming, climbing, etc.)** found that 89% of participants felt that their activity participation was affected by menstruation/PMS. Barriers to participation in adventurous activities included challenges posed by hygiene and waste disposal during menstruation (Prince & Annison 2023).

#### 3.2.2. Interventions to increase exercise participation in relation to the menstrual cycle

- A quasi-experimental study found that **female university students in South Korea** who used **social media-based support** significantly increased their amount of PA and their premenstrual symptoms decreased compared to the control group (Nam & Cha 2020).
- A qualitative workshop in Sweden focused on **designing menstrual technologies** which responded to **adolescents’ premenstrual symptoms** in collaboration with the participants. They found that a toolkit approach to the design of menstrual technologies allowed for pluralist experiences of menstrual cycles (Sondergaard et al. 2021).
- The **Big Sister project** in 11 deprived areas of the UK aimed to empower and support **adolescent girls** to enjoy sport, exercise and PA during puberty. Some of the barriers it aimed to tackle were period poverty and stigma. By collaborating with the “Hey Girls” project (an award-winning “buy one, donate one” social enterprise) they provided free sanitary products for girls in areas of deprivation. Different types of products and information about periods were distributed in women’s toilets/changing rooms at leisure centres, schools, and community projects. Evaluation of The Big Sister project showed positive impact on girls’ engagement in sport and PA with 44% of girls doing a lot more exercise, 64% enjoying taking part, 61% feeling motivated to take part and 58% more confident to take part in sport and exercise (Women in Sport 2023).

#### 3.2.3. Wider barriers to physical activity participation of women, girls, and people who menstruate

- An overview of reviews identified multiple factors that affect **adolescent girls’** participation in PAs. The most common **barriers** were lack of support from peers and/or family, and limited time, while family, peer and teacher support and weight loss were identified as facilitators of PA participation. Identified areas for practice and policy implementation included: inclusive approach to addressing gender norms in the curriculum; training for professionals working with adolescents; environmental change to ensure safety and attractiveness of the PA settings; engagement of multiple stakeholders at local, regional, and national level in incorporating a gender-responsive approach toward PA participation (Duffey et al. 2021).
- A systematic review of qualitative studies identified **adolescent girls’ barriers** to PA participation, such as gender bias in sport, motivation and perceived competence, and competing priorities. Meeting societal expectations, such as peer and adult influences were seen to have both positive and negative aspects regarding PA participation. Community influences were also identified, and rural settings were associated with limited choice of activities. In urban settings while there was a variety of activities, financial constraints often **hindered participation** (Corr et al. 2018).
- A mixed methods systematic review reported that **postpartum women’s barriers** to exercise commonly included tiredness/lack of sleep, lack of time due to domestic chores/responsibilities, and insufficient support from family, friends and other mothers, weather, and breast feeding. Additionally, it was advised that physical therapists and other healthcare professionals should be aware of the unique barriers to exercise amongst postpartum women to provide counselling strategies and meaningful education to this population (Edie et al. 2021).
- A systematic review revealed that **pregnant women** had positive attitudes regarding PA, acknowledging its importance and benefits. **Barriers** were often intrapersonal and included fatigue, lack of time, and pregnancy related discomfort. Additionally, enablers included maternal and foetal health benefits, social support, and pregnancy specific programs. Person-centred strategies using behaviour change techniques should be used to address barriers and translate pregnant women’s positive attitudes into increased PA participation. There was little information found on barriers and enablers for PA in pregnant women with gestational diabetes (Harrison et al. 2018).
- A systematic review reported **factors** associated with **adolescent girls’ sport participation** including personal, peer, family, socioeconomic, and other circumstances. Personal factors, including self-perceptions and desirable personal outcomes related to sport, were most frequently associated with participation (Hopkins et al. 2022).
- A systematic review identified **barriers** to PA for **young adult women** that included factors such as time, body image, societal beauty standards, family duty, social support, religious and cultural norms, safety issues, facilities and physical environment (Peng et al. 2023).
- A systematic review identified multiple factors that influenced **midlife immigrant women’s PA participation**. These included individual, familial and community factors. Individual factors related to health status, limited time, lack of motivation and proficiency in life skills, such as language proficiency, driving, and PA knowledge. Familial factors consisted of family support and seasonality (as in seasonal celebrations), while community factors related to social support and physical environment (Zou et al. 2021).

#### 3.2.4. Interventions (unrelated to managing the menstrual cycle) to support PA participation of women, girls and people who menstruate

- A systematic review found that **PA interventions** may encourage **adolescent girls and young adults (11-25 years)** to try new sports. However, there was limited evidence of sustained participation. Recommended strategies included consultation with girls, implementation of peer-leaders and friendship group elements, early intervention, and considerations for setting (Allison et al. 2017).
- A systematic review with a meta-analysis conducted by AlSwayied et al. (2022) showed that there was a small to moderate increase of 61.38 weekly minutes in moderate to vigorous PA in **midlife women undergoing menopause** following **mobile based PA interventions**. Positive improvements were also found in menopause-related outcomes, such as weight reduction, anxiety management, sleep quality, and menopause-related quality of life. Participants perceived mobile PA interventions to be acceptable and helpful.
- A systematic review with a meta-analysis indicated that **PA interventions for pre-adolescent girls** had a small statistically significant effect in increasing participation. Subgroup analysis indicated that interventions that were **educational or multicomponent**, or those aimed at **girls only** were more effective than others (Biddle et al. 2014).
- A systematic review suggested that the most effective interventions to promote PA among **young and adolescent girls** were those that were **school-based and multicomponent with PE tailored to girls’ unique needs**. Additionally, it showed influence of peers more promising than family support (Camacho-Minano et al. 2011).
- A systematic review and meta-analysis of cross-sectional studies suggested that **social support** is not a strong predictor of PA in **adolescent girls**. However, support from parents and friends may have a role in changing girls’ PA behaviour (Laird et al. 2016).
- A systematic review and meta-analysis found that **interventions that target the intersection of body image and movement** for **girls and women** led to a small statistically significant improvement in body image at post-test. However, long-term effect detected at follow-up was not significant. Additionally, while body image and movement focused interventions had an effect on fitness outcomes, they were found to have no statistically significant effect on movement behaviour (Matheson et al. 2023).
- A systematic review by National Institute for Health and Care Excellence (NICE) (2008) identified school and primary care based interventions that promoted PA for **adolescent girls**. These interventions consisted of mediated (delivered via computer, phone or printed material) counselling and educational elements. Findings indicate that **school-based interventions outside of PE targeting PA behaviour only** could have a moderate-to-large effect.
- A systematic review found that **telehealth interventions** in **postpartum women** could increase moderate to vigorous PA (Turner et al. 2023). While the majority of included studies showed an effect (n=10), some studies had no significant impact. Findings indicate that video consultations might be most effective.

#### 3.2.5. The effects of physical activity on dysmenorrhea

- A systematic review with meta-analysis found low quality evidence that **exercise for 45-60 minutes three times or more weekly** may provide a **significant reduction in menstrual pain intensity**, and therefore could be used solely or in conjunction with other modalities such as non-steroidal anti-inflammatory drugs (Armour et al. 2019a).
- A systematic review with meta-analysis suggested that **self-care and lifestyle interventions**, such as self-directed acupressure, exercise, and heat all reduced menstrual pain symptoms. **Exercise had the largest effect** and alongside heat was more effective at reducing menstrual pain symptoms than analgesics (Armour et al. 2019b).
- A systematic review found moderate to low certainty evidence that **PA could effectively reduce menstrual pain intensity and duration**. However, there is a need for high quality trials (Matthewman et al. 2018).

#### 3.2.6. The effects of physical activity on premenstrual syndrome

- A systematic review with meta-analysis suggests that **exercise interventions could reduce overall, psychological, physical, and behavioural symptoms** of PMS (Pearce et al. 2020). However, due to heterogeneity of included studies and high risk of bias, uncertainty remains.
- A systematic review found that **exercise improved premenstrual symptoms in women** including physical and psychological symptoms, such as pain, constipation, breast sensitivity, anxiety and anger. (Saglam & Orsal 2020).

#### 3.2.7. The menstrual cycle and exercise performance

- An overview of reviews found largely **inconsistent evidence** of marked differences in strength, exercise performance, and hypertrophy throughout menstrual cycle phases (Colenso-Semple et al. 2023).
- A scoping review suggested that **oestrogen and progesterone is responsible for differences in athletic performance throughout the menstrual cycle** (Bernstein & Behringer 2023). However, the evidence base mainly consists of animal and in vitro research, and more high quality human studies using cycle monitoring methodology is needed.
- A systematic review with meta-analysis found that **exercise performance had a very small reduction during the early follicular phase** of the menstrual cycle, compared to all other phases. (McNulty et al. 2020).
- A systematic review found a **variable association between menstrual cycle and elite athletes’ performance-related outcomes**, such as endurance or power resistance, ligament stiffness, decision making skills, psychology, or competitiveness. (Meignié et al. 2021).
- A systematic review with meta-analysis suggested that the **early follicular phase is unfavourable for all maximal strength performance**. Isometric strength peaks and dynamic strength is optimal in the late follicular phase, while isokinetic strength is the highest during ovulation (Niering et al. 2024).
- A systematic review and meta-analysis found that **menstrual cycle phases did not impact rating of perceived exertion** during exercise (Prado et al. 2023). However, further research is needed on the topic.

### 3.3. Areas of uncertainty

Remaining uncertainties include:

- There seems to be a lack of secondary and primary evidence on interventions that could support the PA (including exercise and sport) participation of women, girls, and people who menstruate in relation to the menstrual cycle.

## 4. NEXT STEPS

While there seems to be a lack of evidence regarding interventions that could support the PA (including exercise and sport) participation of women, girls and people who menstruate in relation to the menstrual cycle, a wealth of secondary research evidence is available for PA promotion that is unrelated to the menstrual cycle. Following the intermediary stakeholder meeting, it was decided to conduct a rapid overview of reviews on the effectiveness of interventions that support women, girls, and people who menstruate to participate in exercise/sport/physical activity. The protocol is registered on Open Science Framework: https://doi.org/10.17605/OSF.IO/XTYCW

## Data Availability

All data produced in the present study are available upon reasonable request to the authors

## Abbreviations

CI: Confidence interval
GP: General practitioner
GRADE: Grading of Recommendations, Assessment, Development, and Evaluations
MC: Menstrual cycle
MVPA: Moderate-to-vigorous physical activity
NHS: National Health Service
NICE: National Institute for Health and Care Excellence
NSAID: Non-steroidal anti-inflammatory drugs
OoR: Overview of reviews
PA: Physical activity
PE: Physical education
PMS: Premenstrual syndrome
RPE: Rating of perceived exertion
ScR: Scoping review
SMD: Standardised mean difference
SR: Systematic review
SUCRA: Surface Under the Cumulative Ranking curve
VAS: Visual analogue scale

## 6. RAPID EVIDENCE SUMMARY METHODS

A list of the resources searched during this Rapid Evidence Summary is provided within Appendix 1. Searches were limited to English-language publications and did not include searches for primary research studies if secondary research relevant to the question was found. Search hits were screened for relevance by a single reviewer.

Priority was given to robust evidence synthesis using minimum standards (systematic search, study selection, quality assessment, appropriate synthesis). The literature identified was not formally quality assessed. The included literature may vary considerably in quality and the degree of such variation will be investigated during the rapid review work (focusing on interventions supporting women, girls, and people who menstruate to participate in exercise/sport/physical activity) which follows on. Citation, recency, evidence type, document status and key findings were tabulated for all relevant research identified in this process.

As secondary research evidence was limited on the topic of interventions/innovations that could support/encourage the PA participation (including exercise and sport) of women, girls, and people who menstruate in relation to the menstrual cycle, targeted searches for primary studies and secondary research in other PA topics were conducted to inform options for further work.

|  |  |
| --- | --- |
| **Date of Search** | March 2024 |
| **Search Concepts Used** | 1. Menstruation AND Exercise AND participation<br>2. Menstruation AND Exercise AND barriers<br>3. Women, girls, female AND Exercise AND barriers<br>4. Exercise performance AND menstruation AND systematic reviews<br>5. Women, girls, female AND Exercise AND participation |
| **Search Completed by** | Cardiff Evidence Synthesis Collaborative |

## 7. EVIDENCE

**Table 1.** Summary of review evidence identified.

| **Evidence type** | **Total identified** | **Comments** |
| --- | --- | --- |
| Overviews of reviews | 2 |  |
| Systematic reviews | 25 |  |
| Scoping reviews | 1 |  |
| Organisational reports | 7 |  |
| Primary Studies | 7 |  |

A more detailed summary of included evidence can be found in Table 2, Table 3 and Table 4.

**Table 2:**
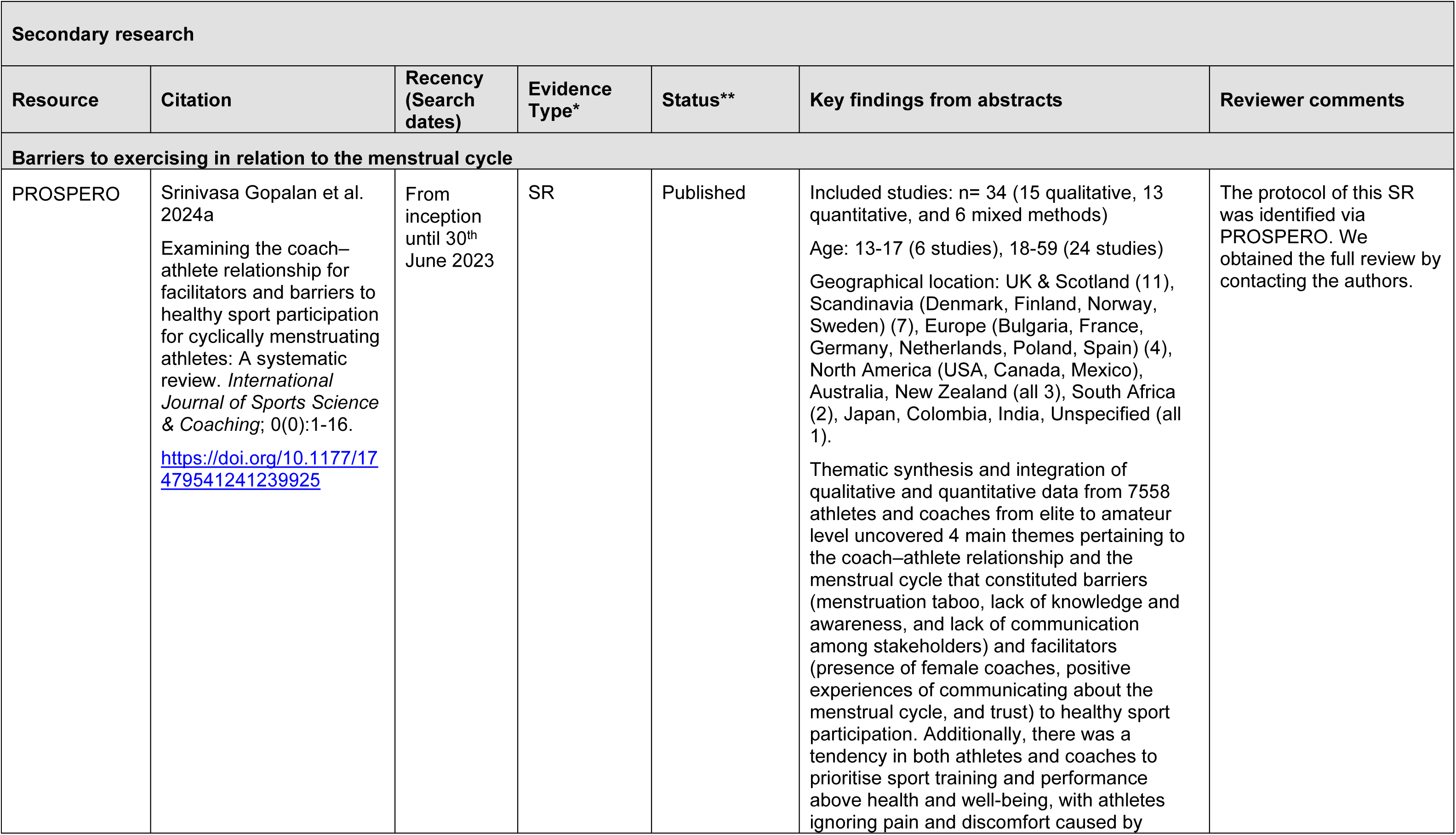
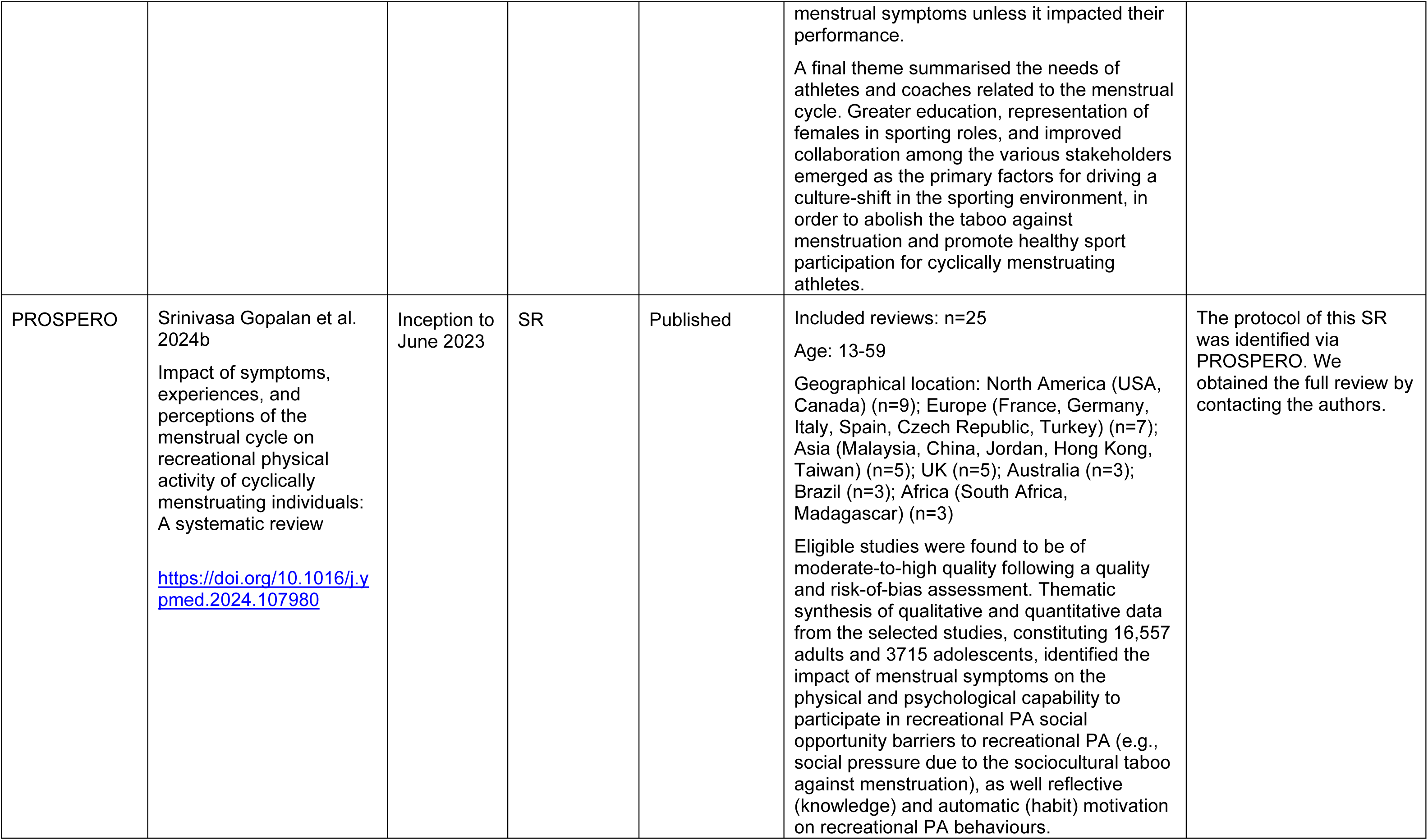
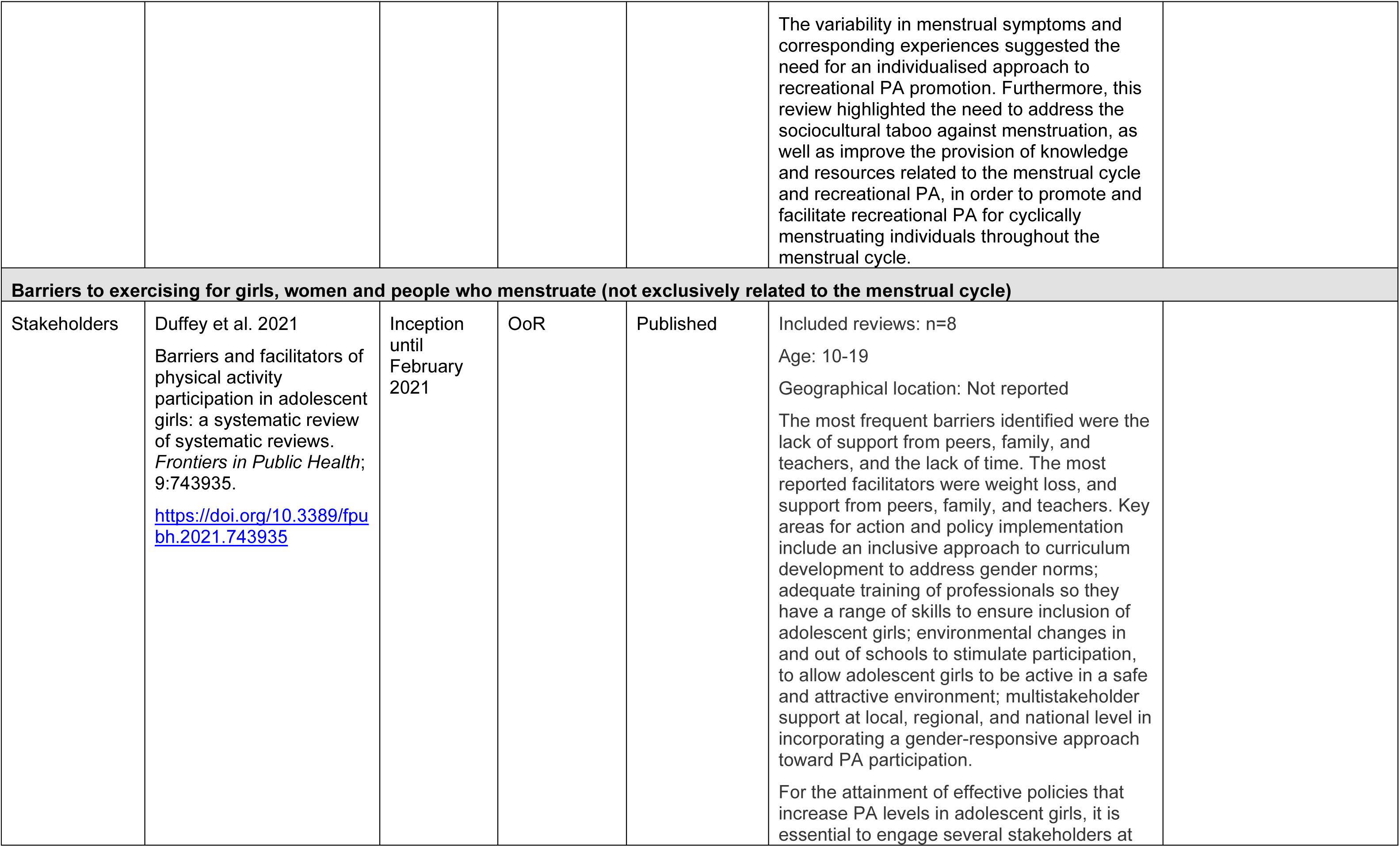
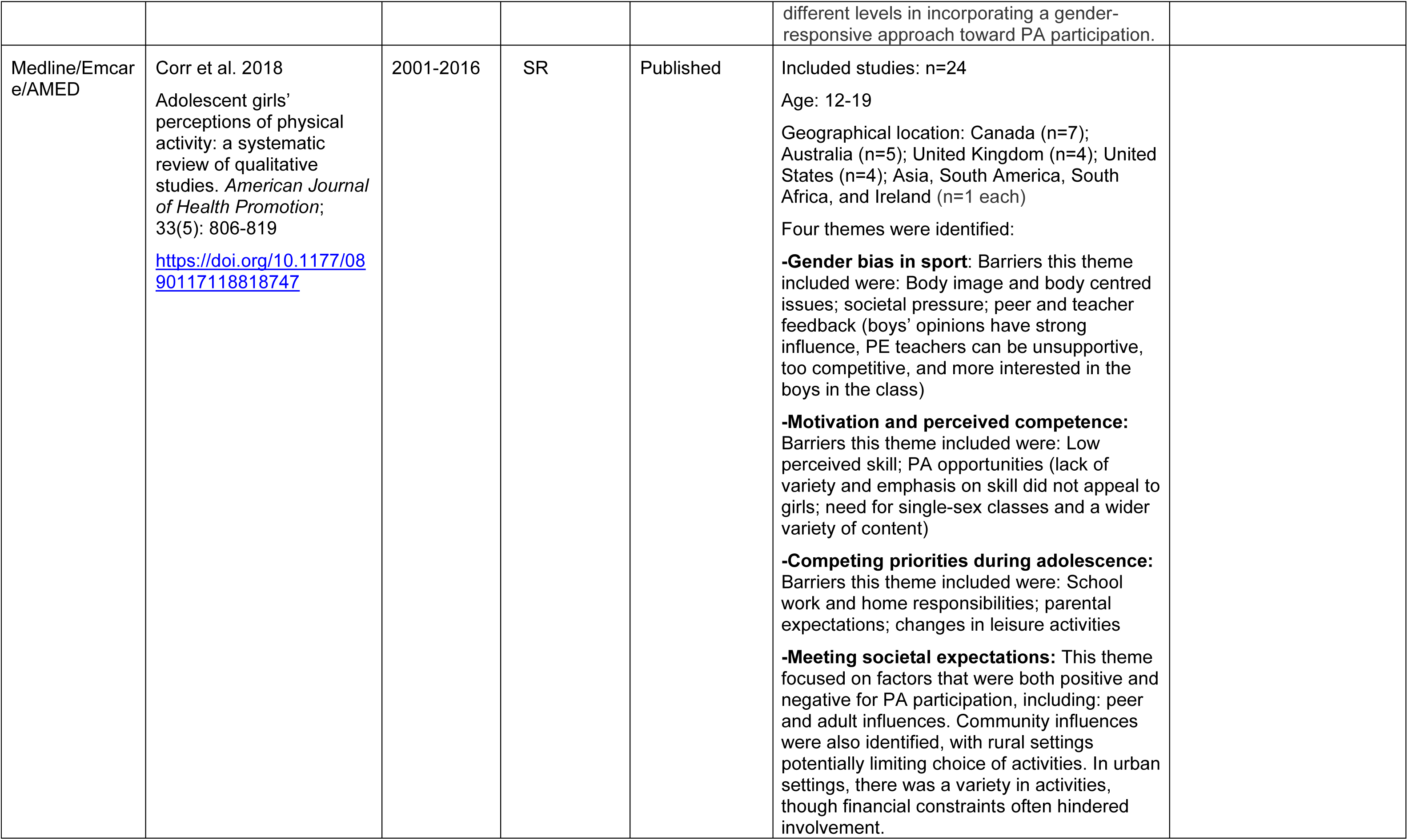
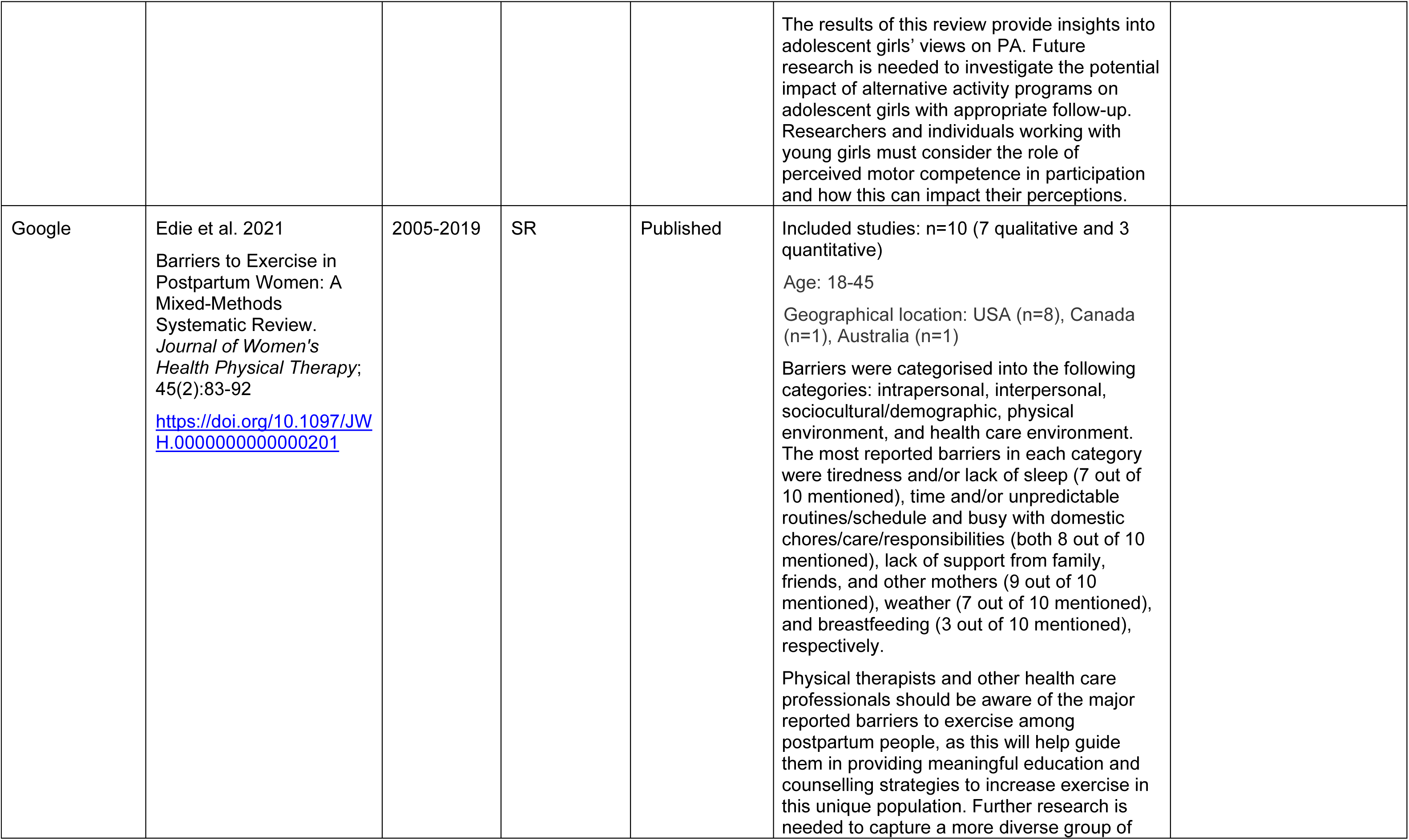
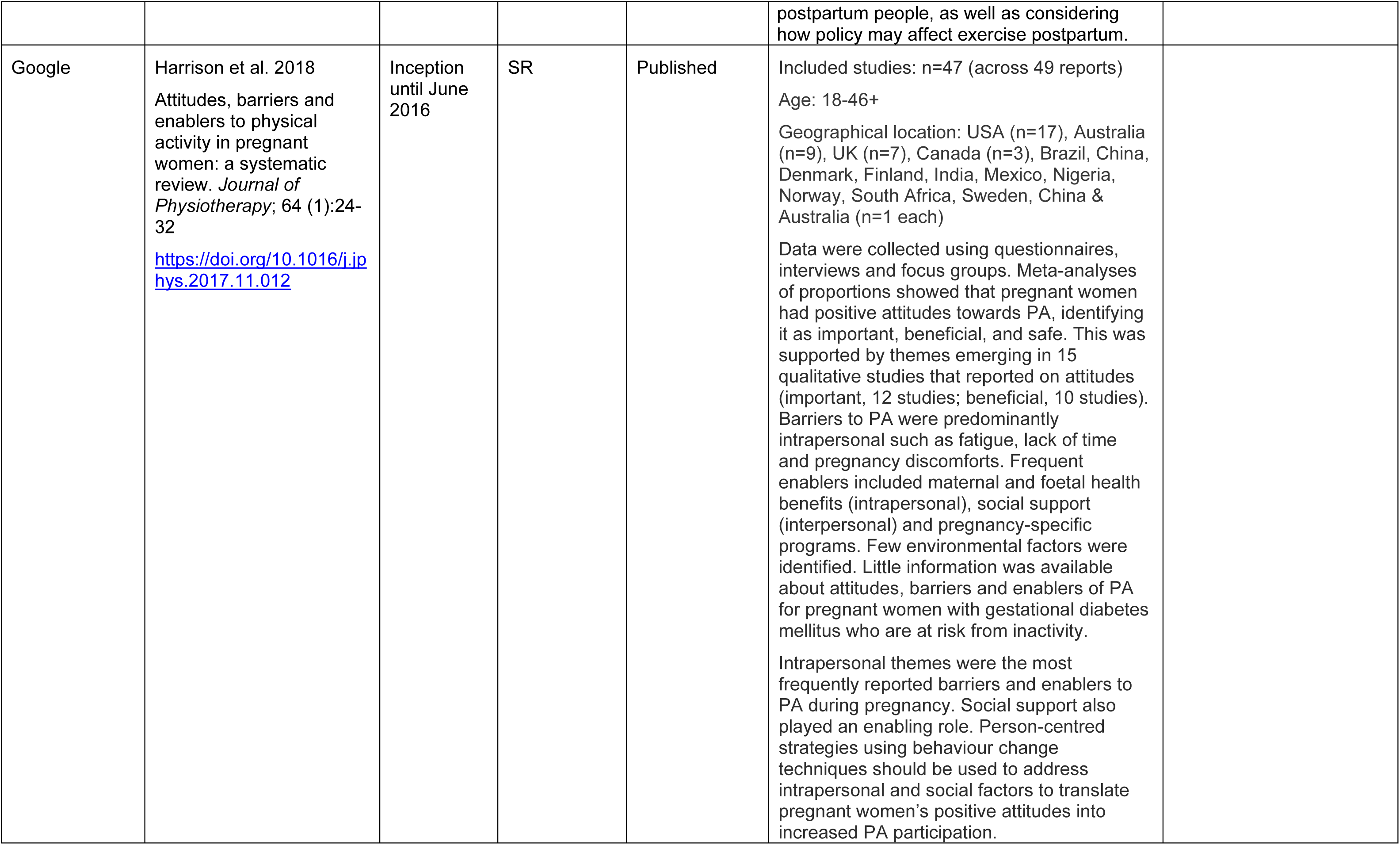
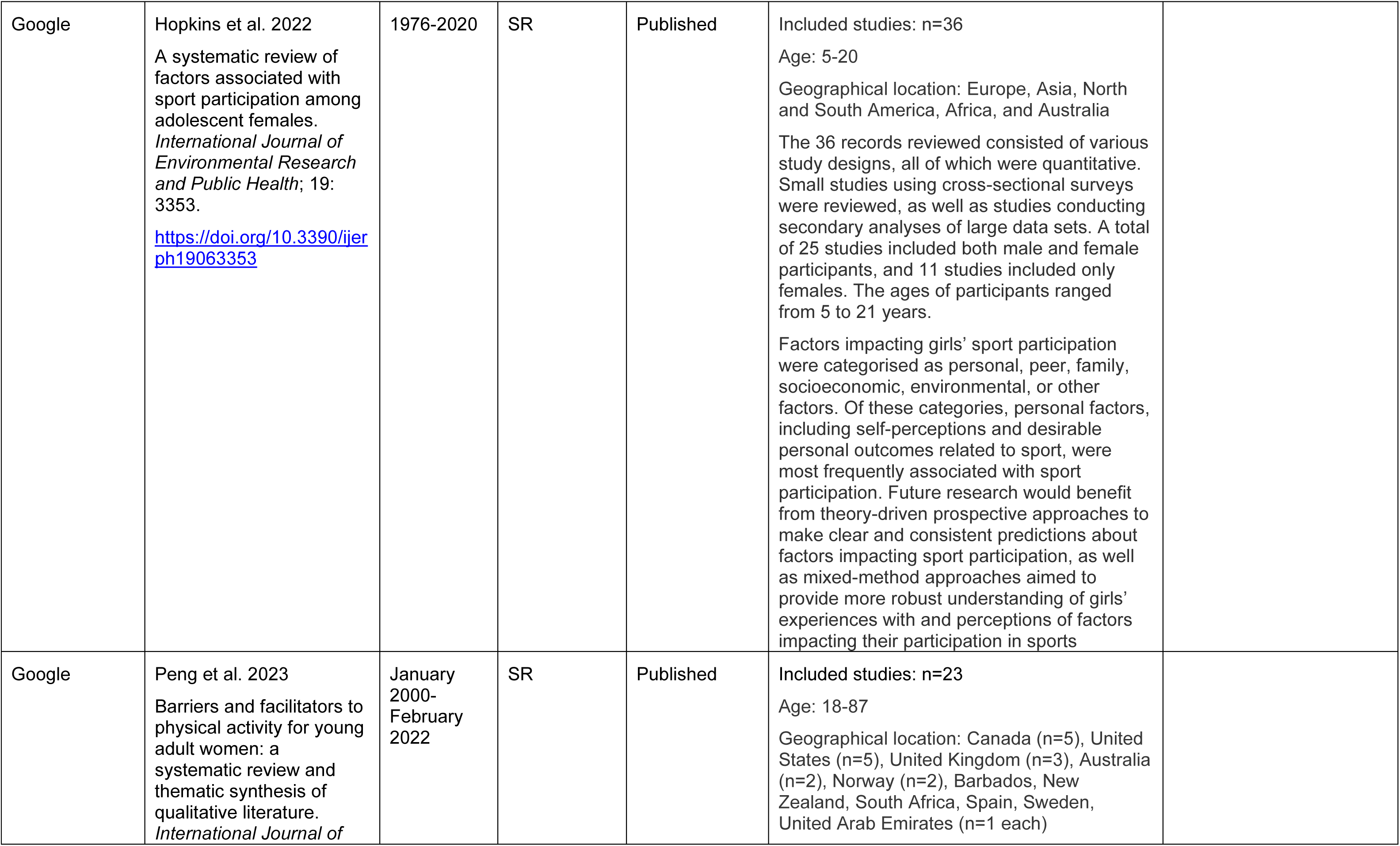
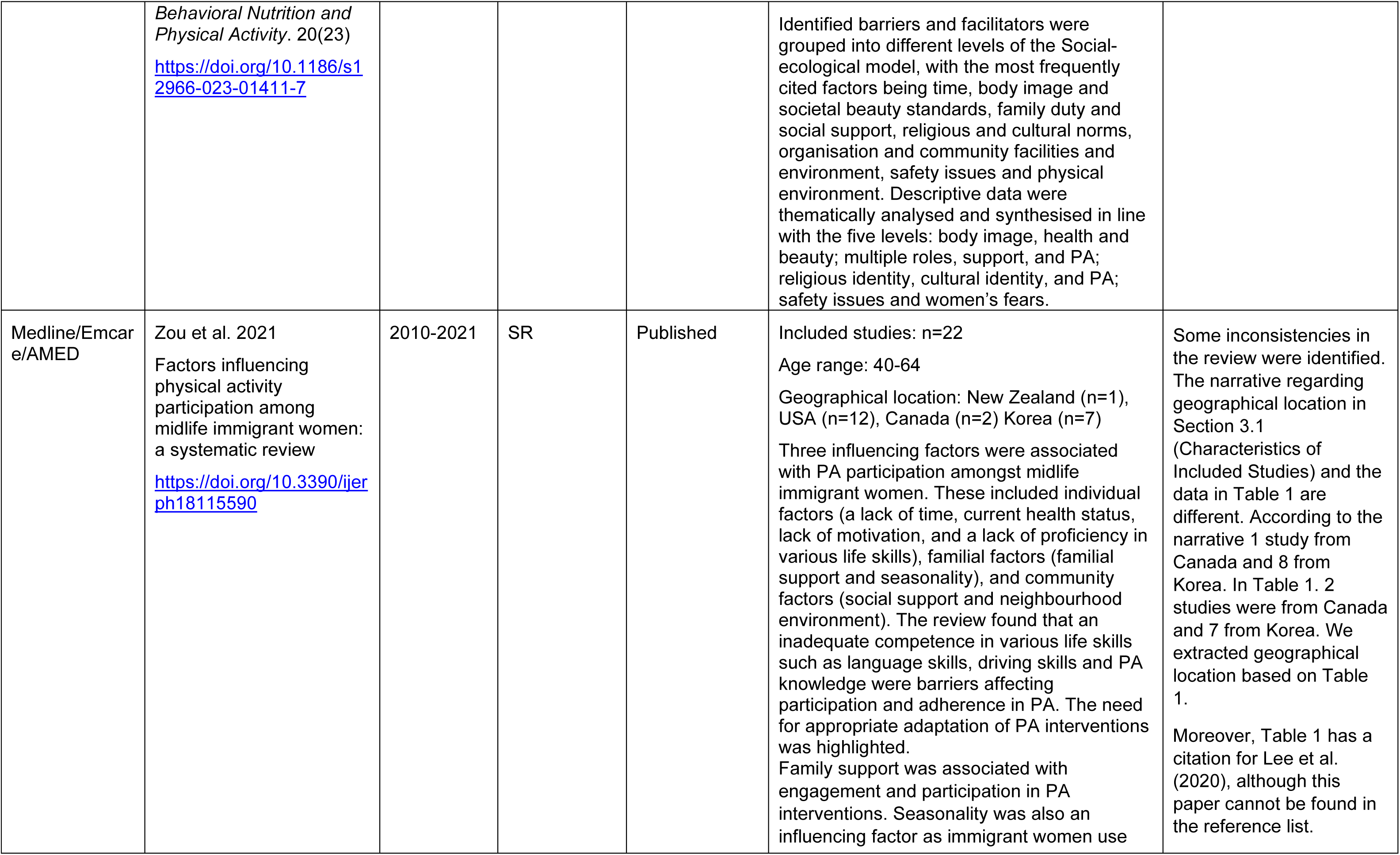
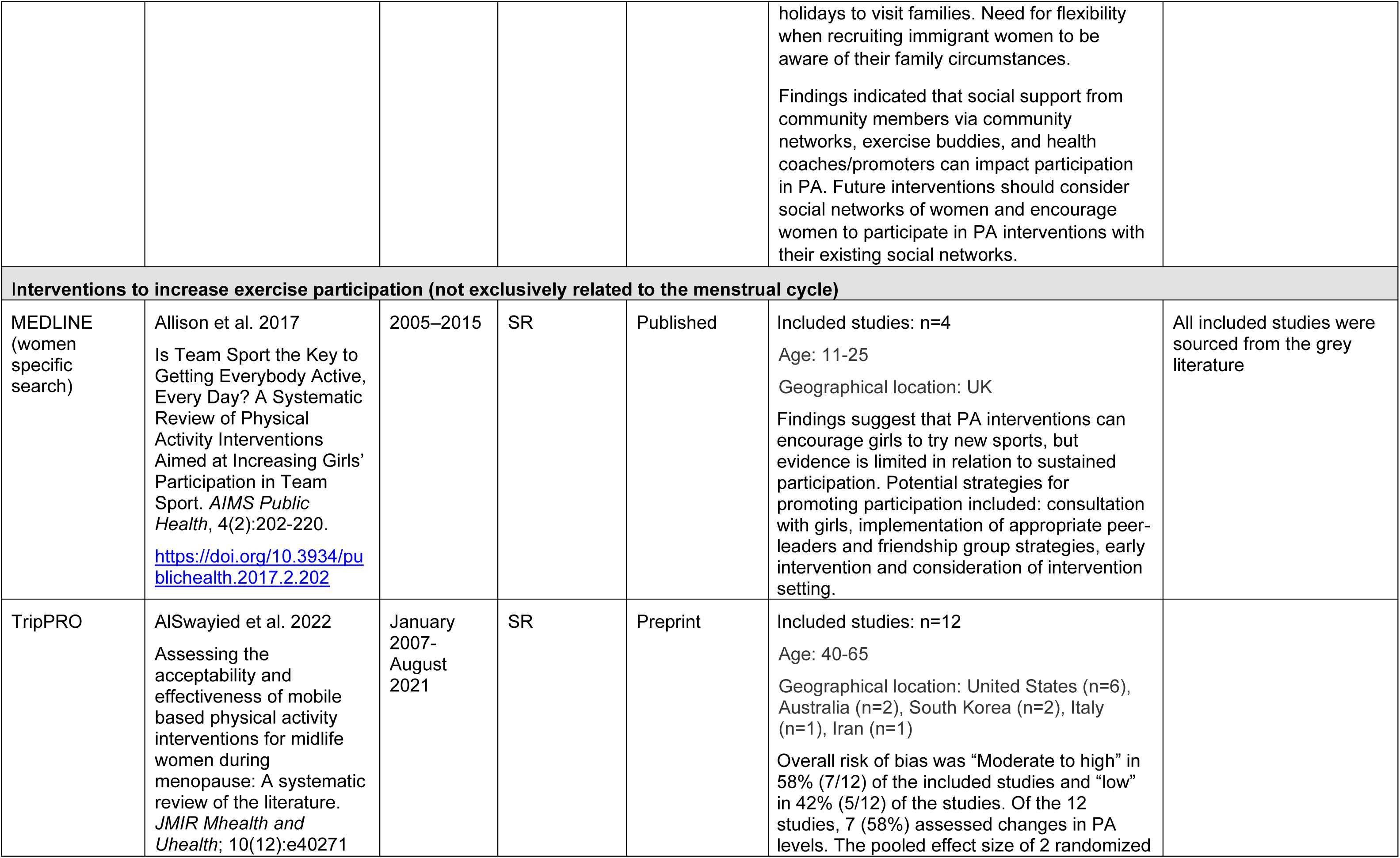
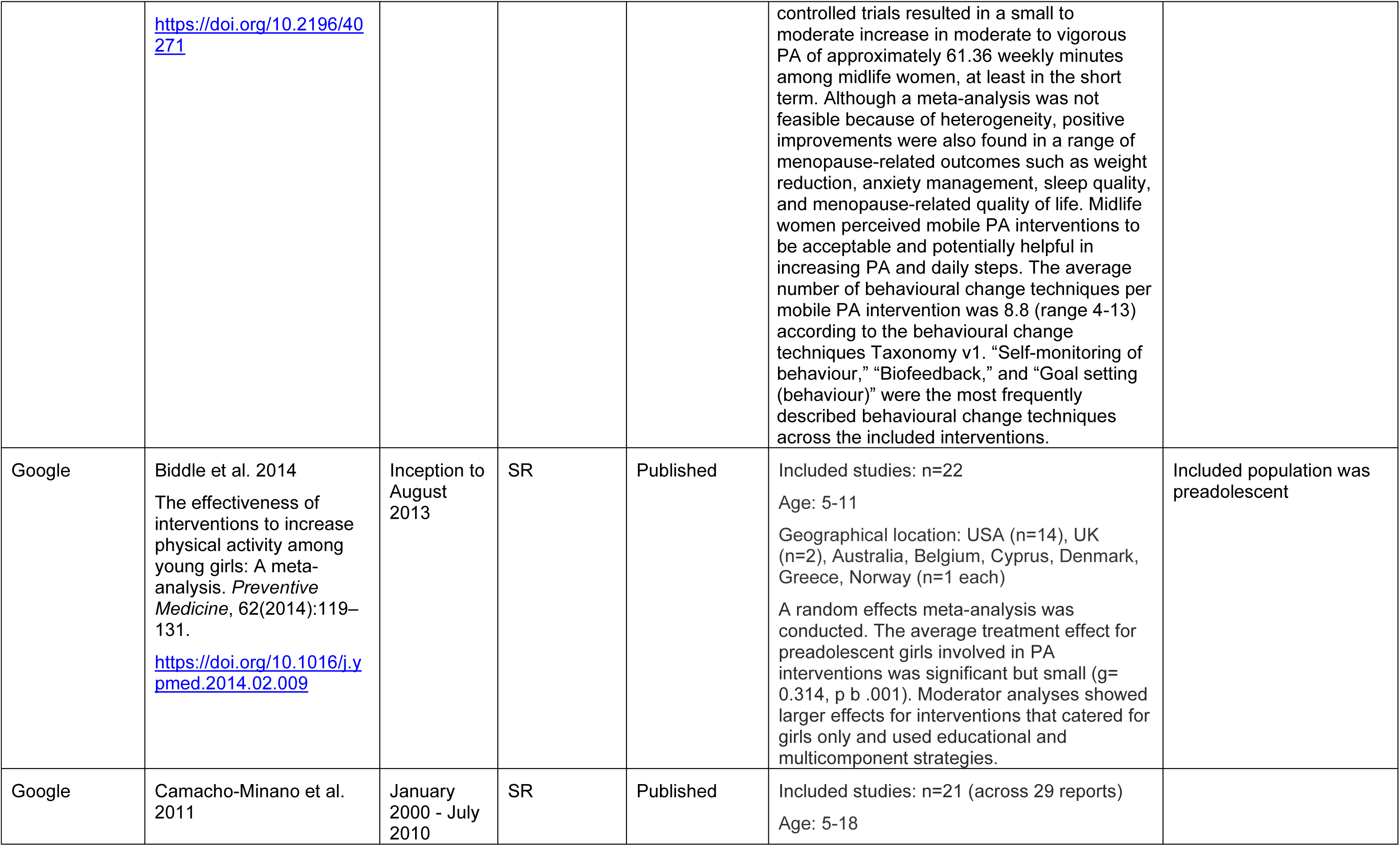
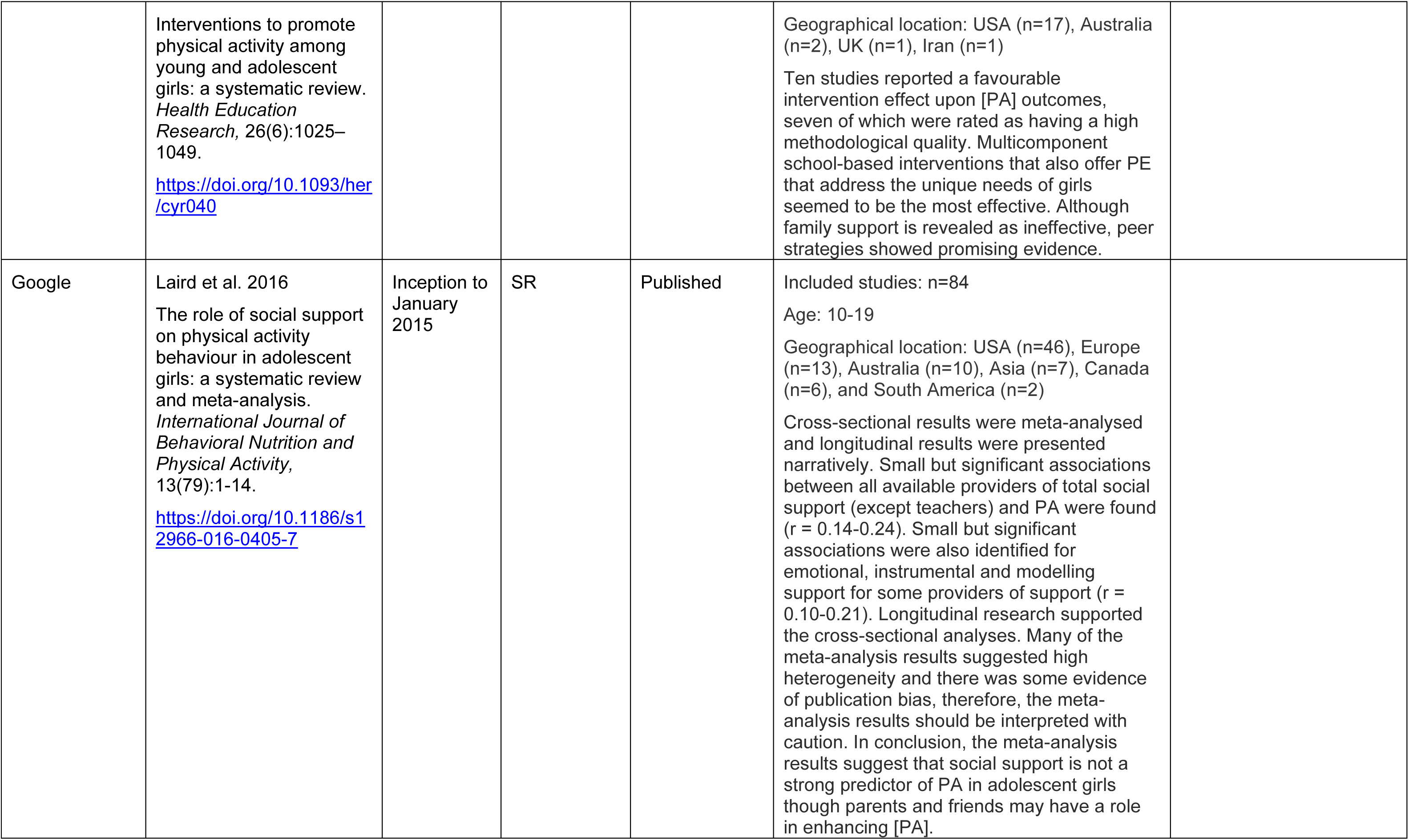
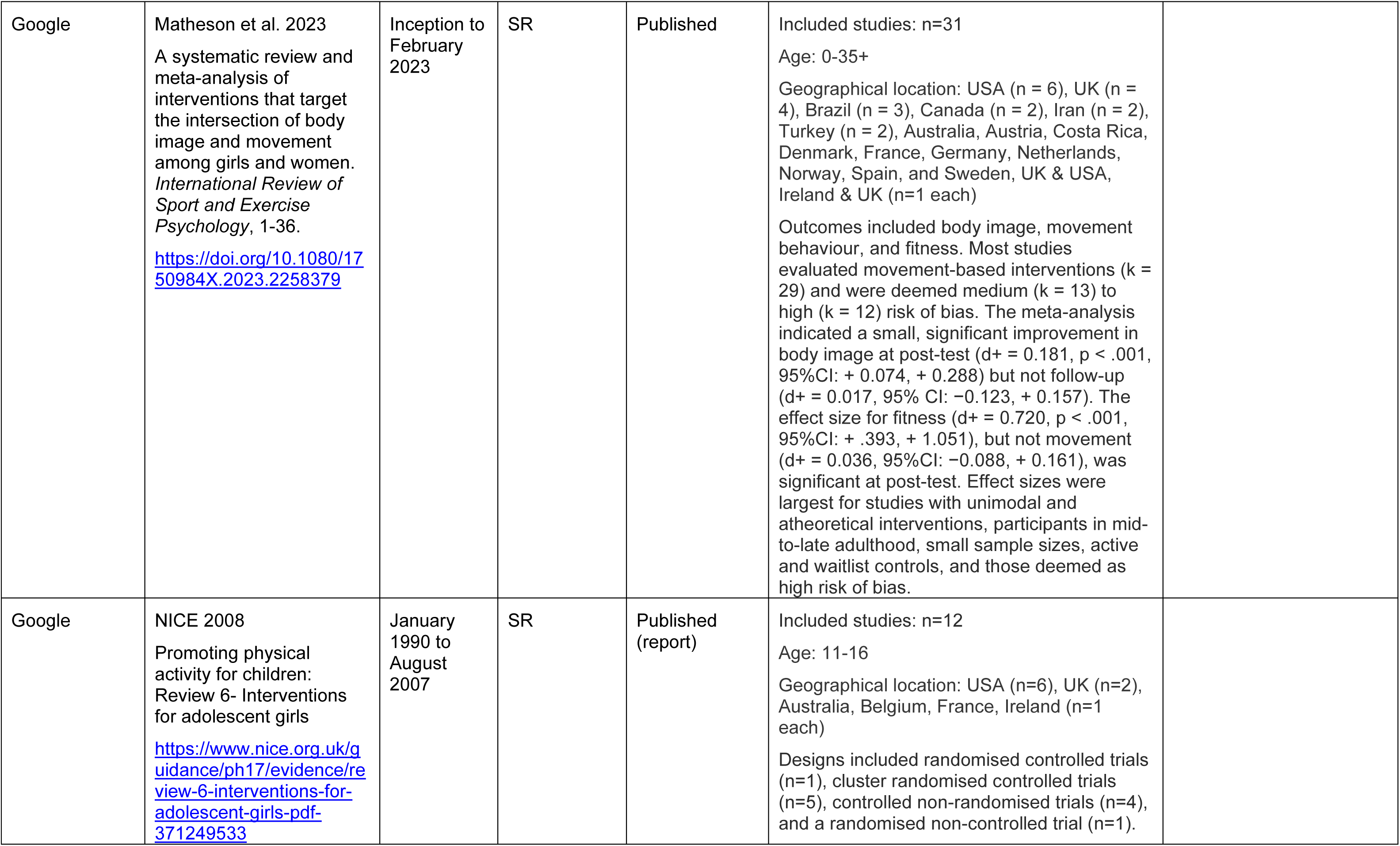
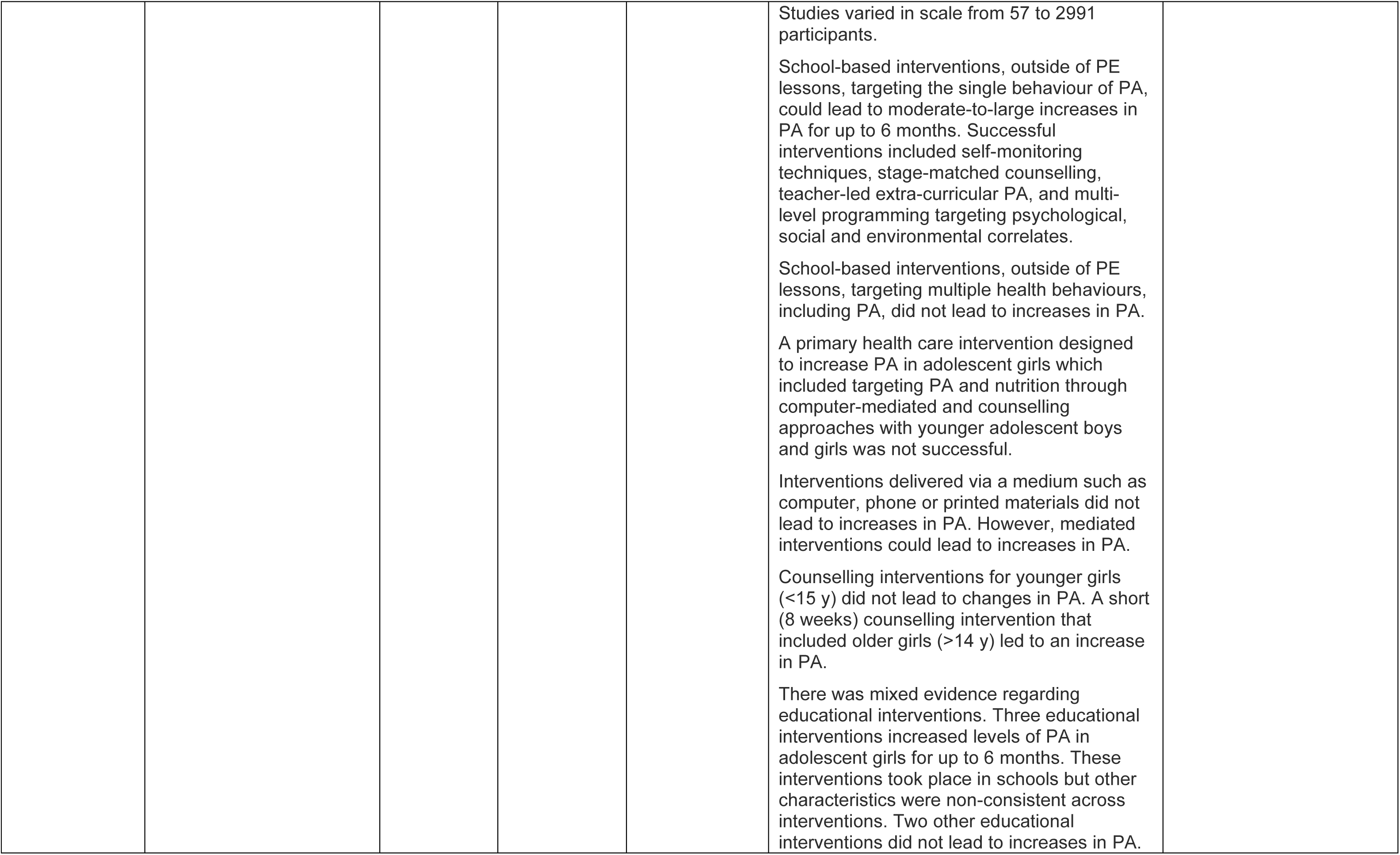
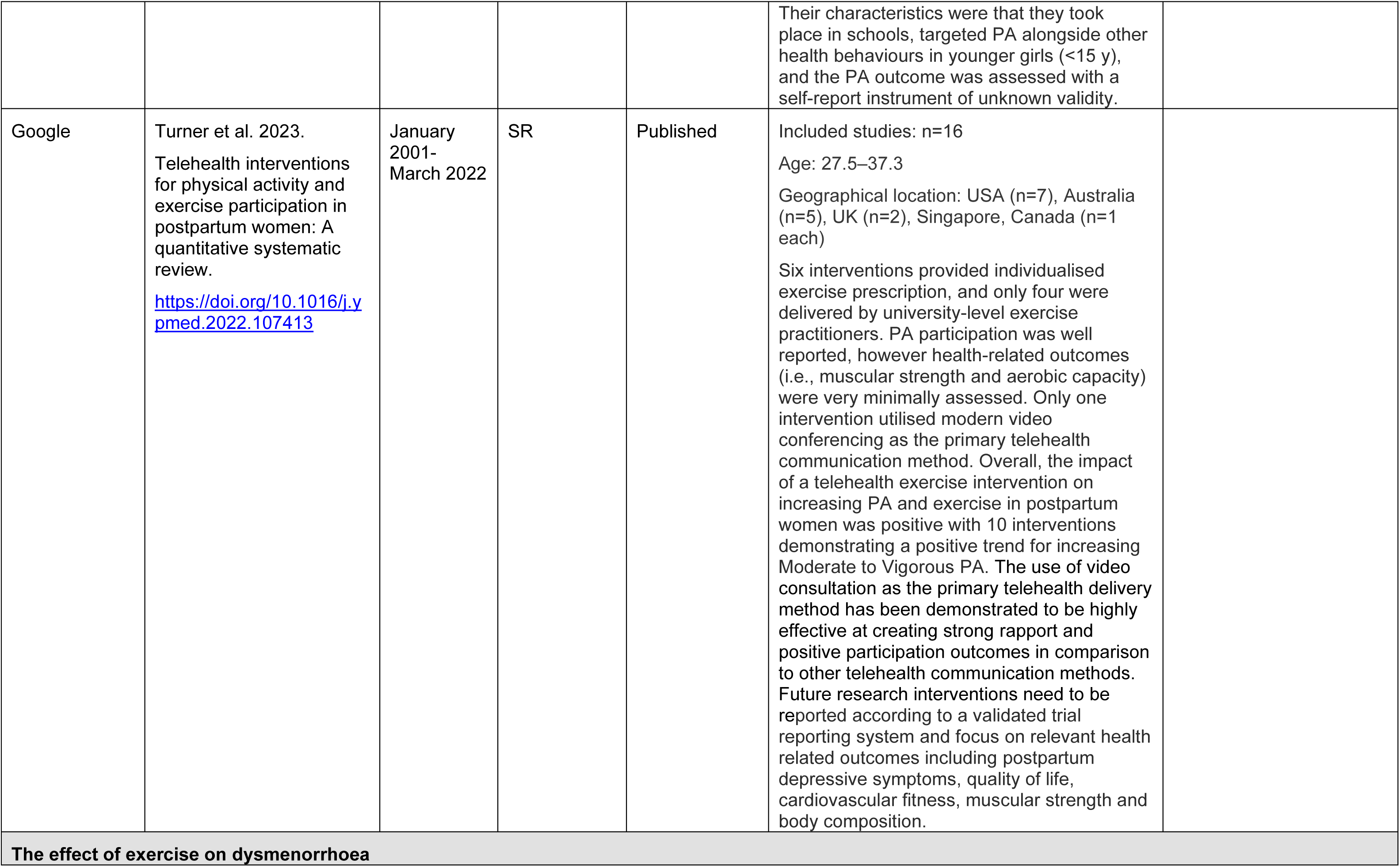
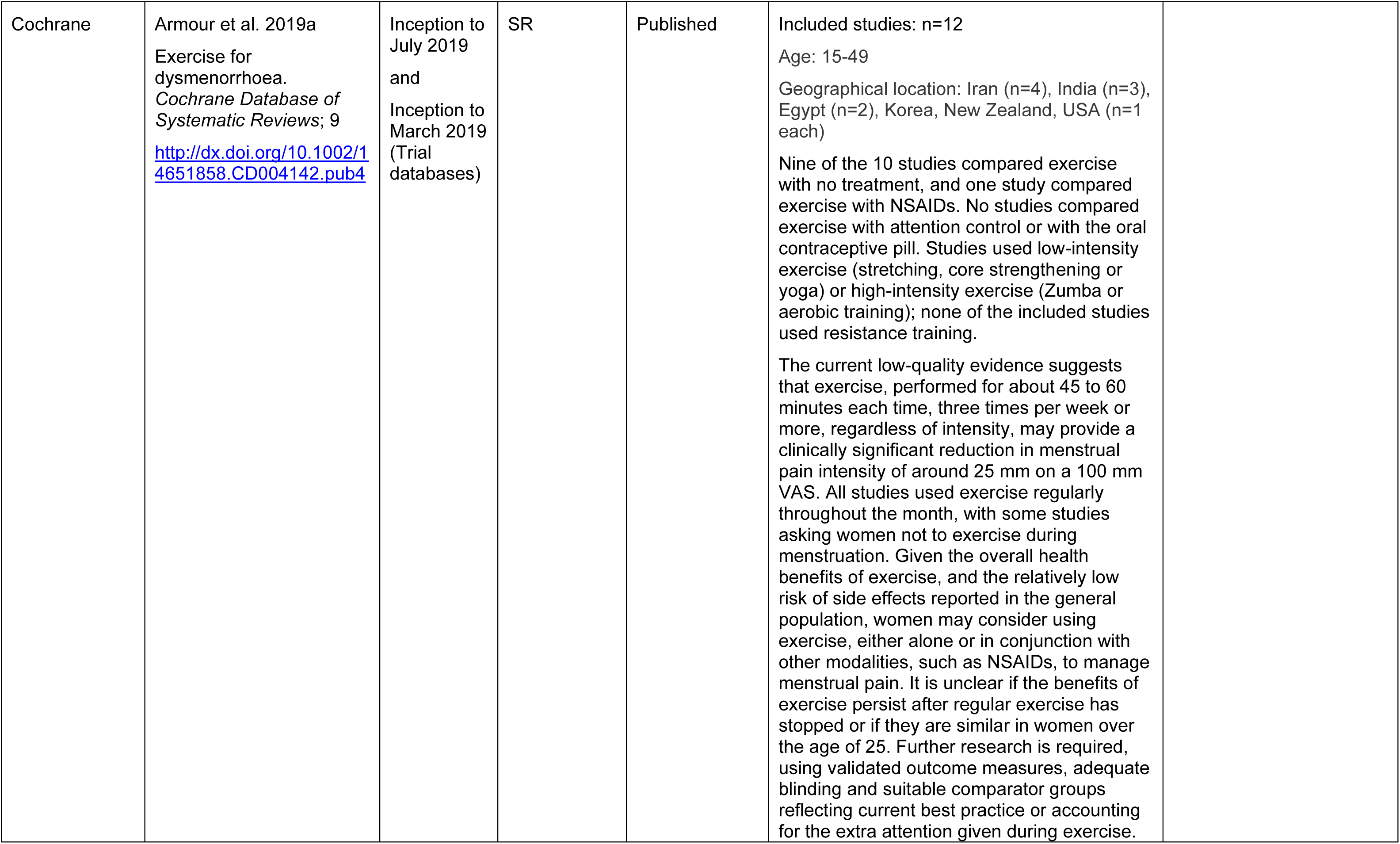
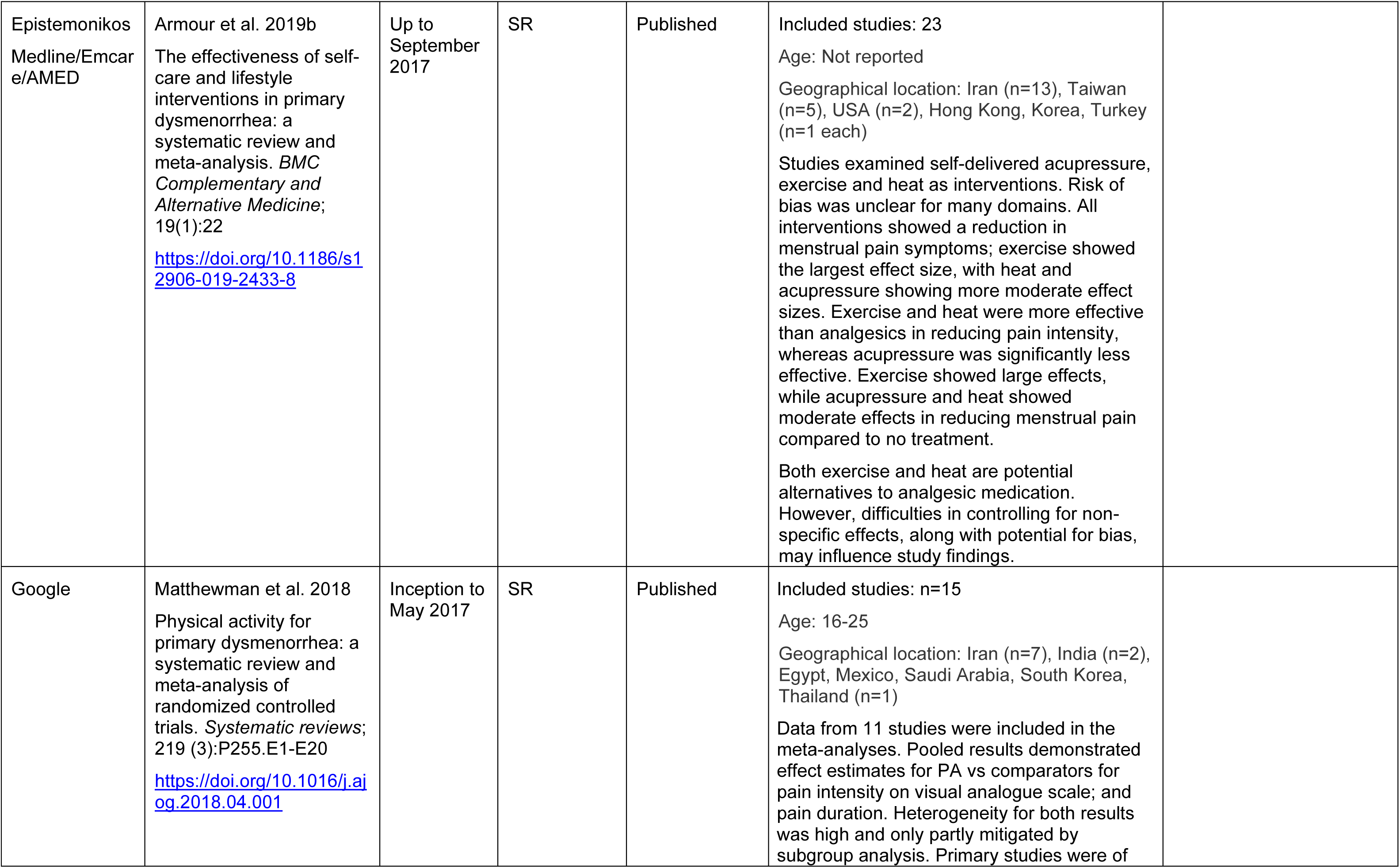
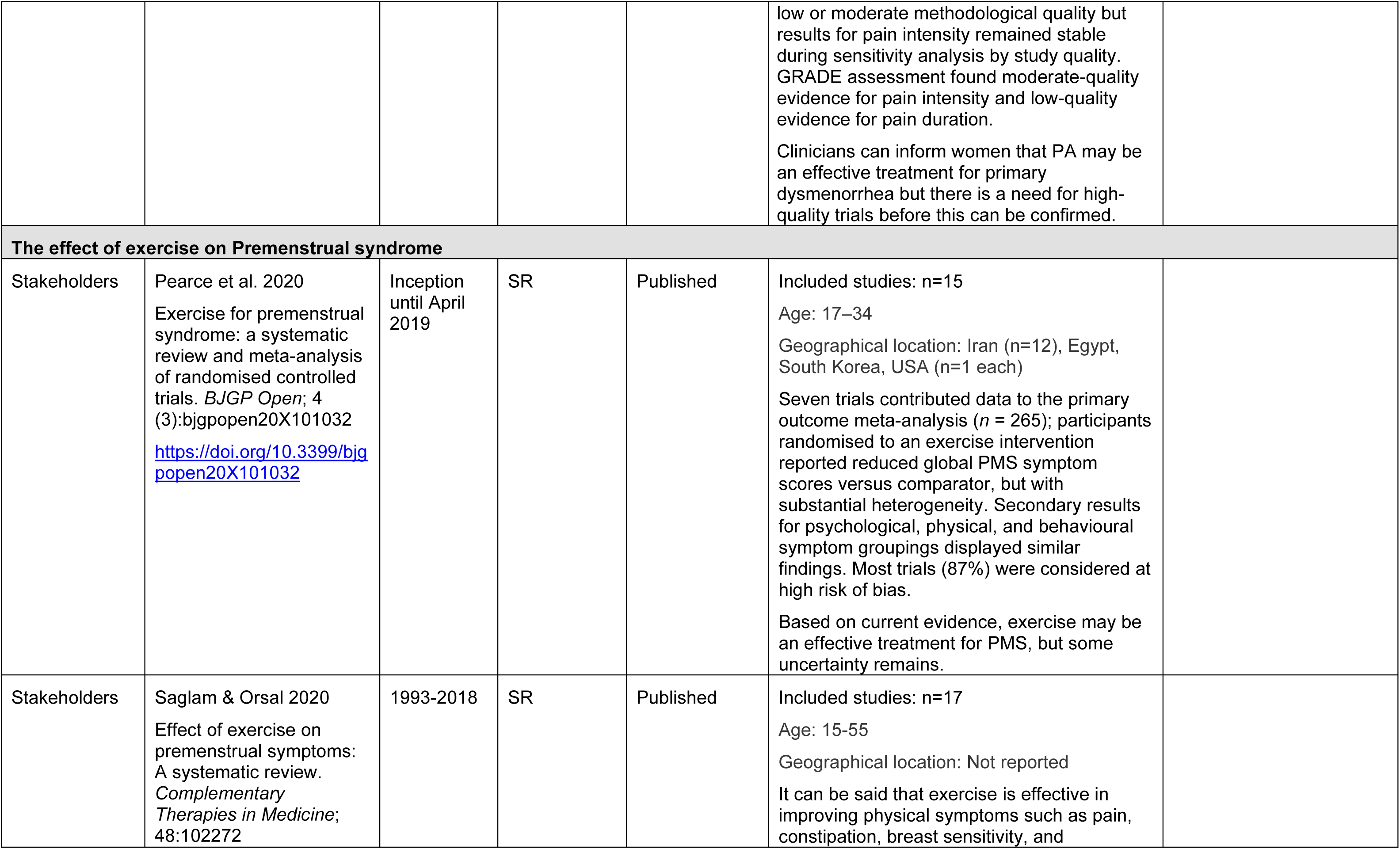
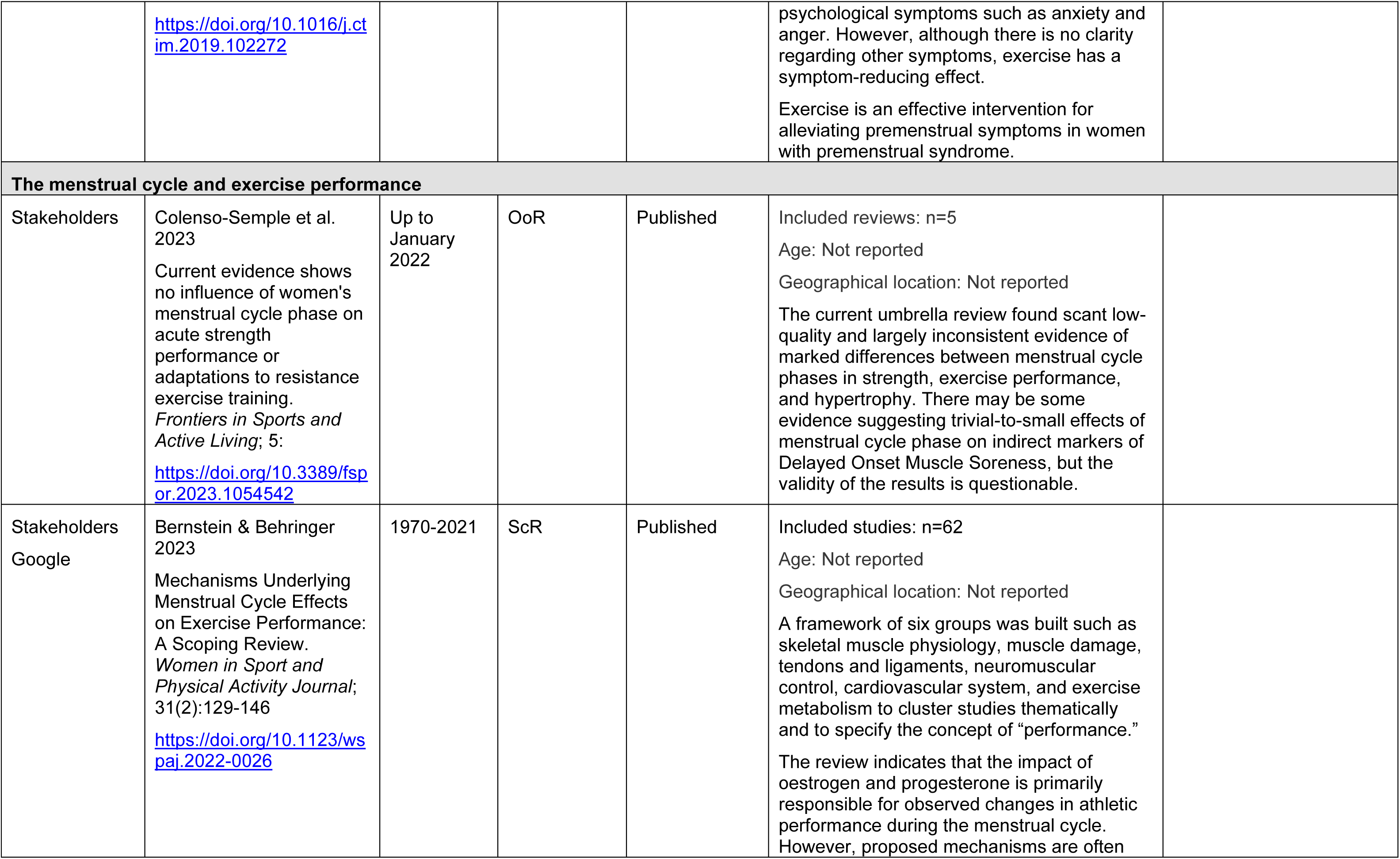
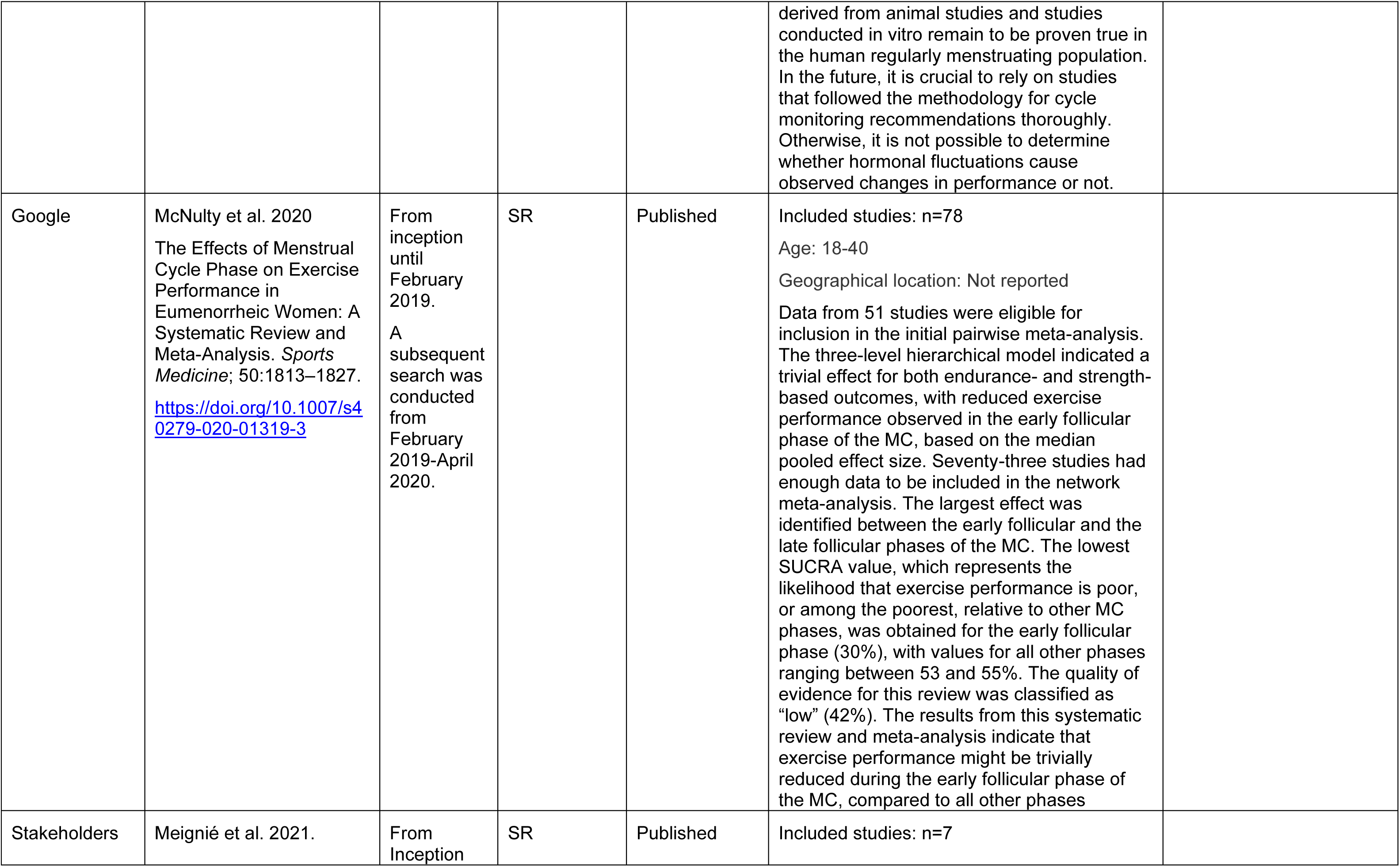
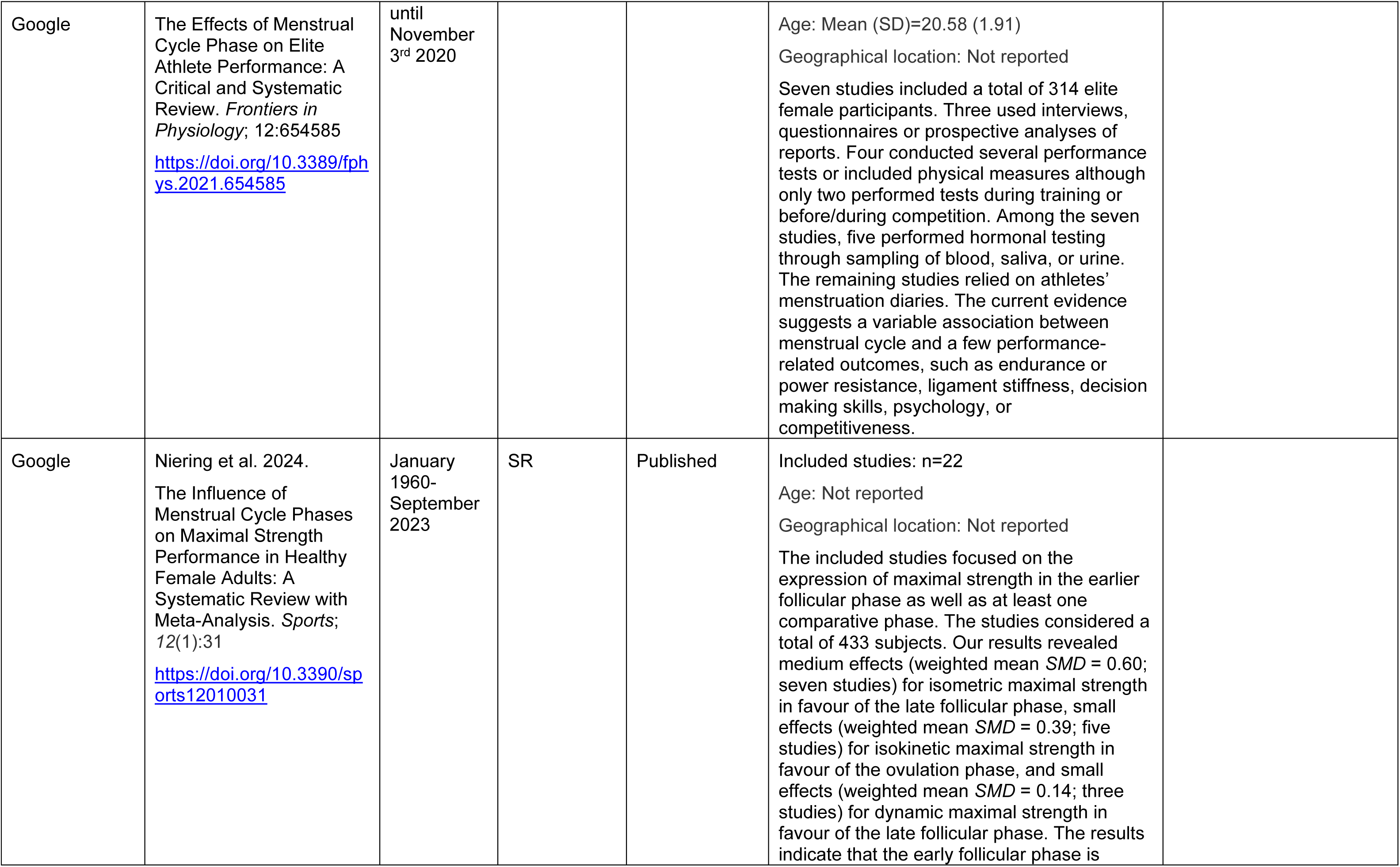
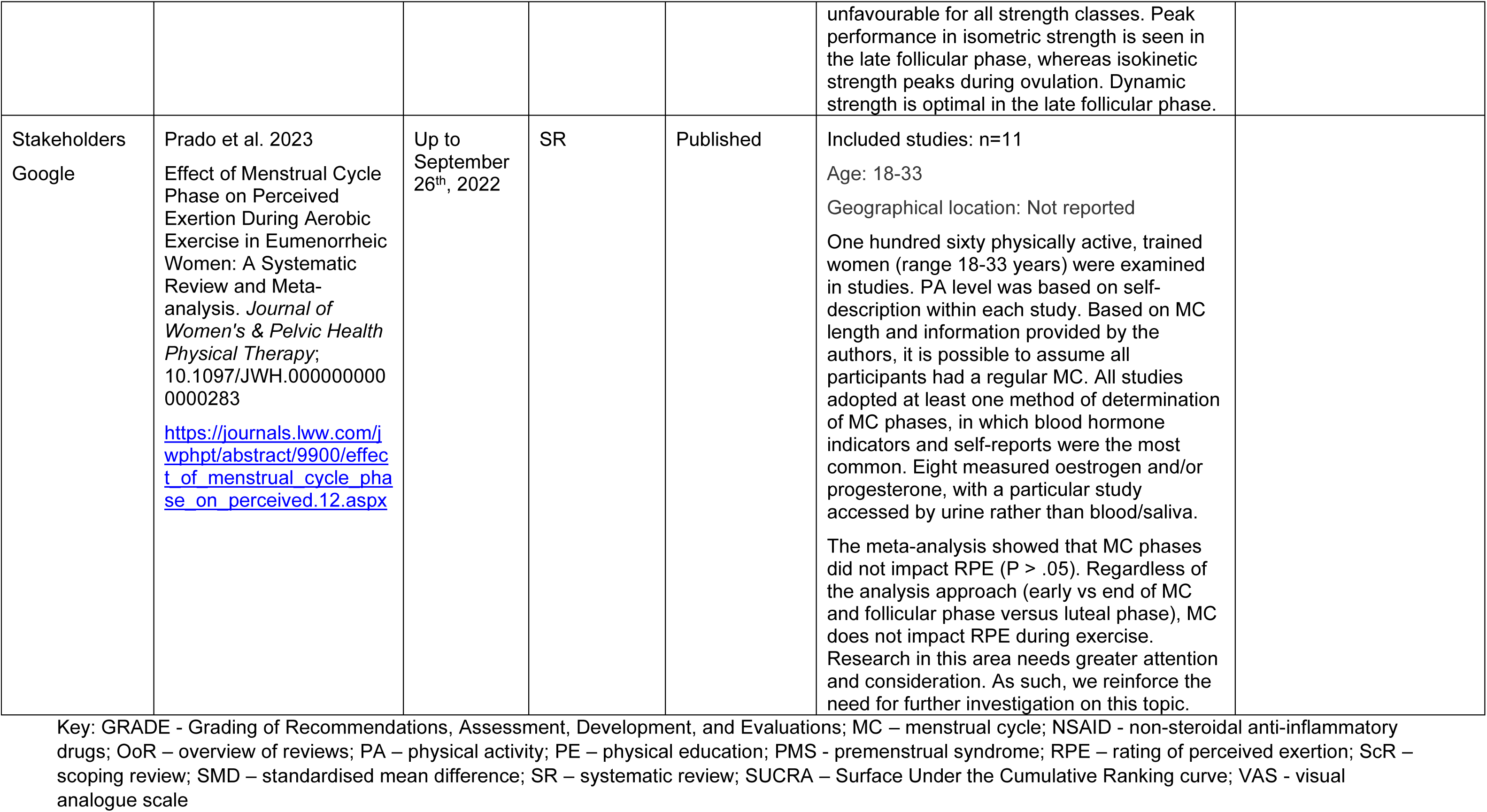
Summary of included secondary evidence.

**Table 3:**
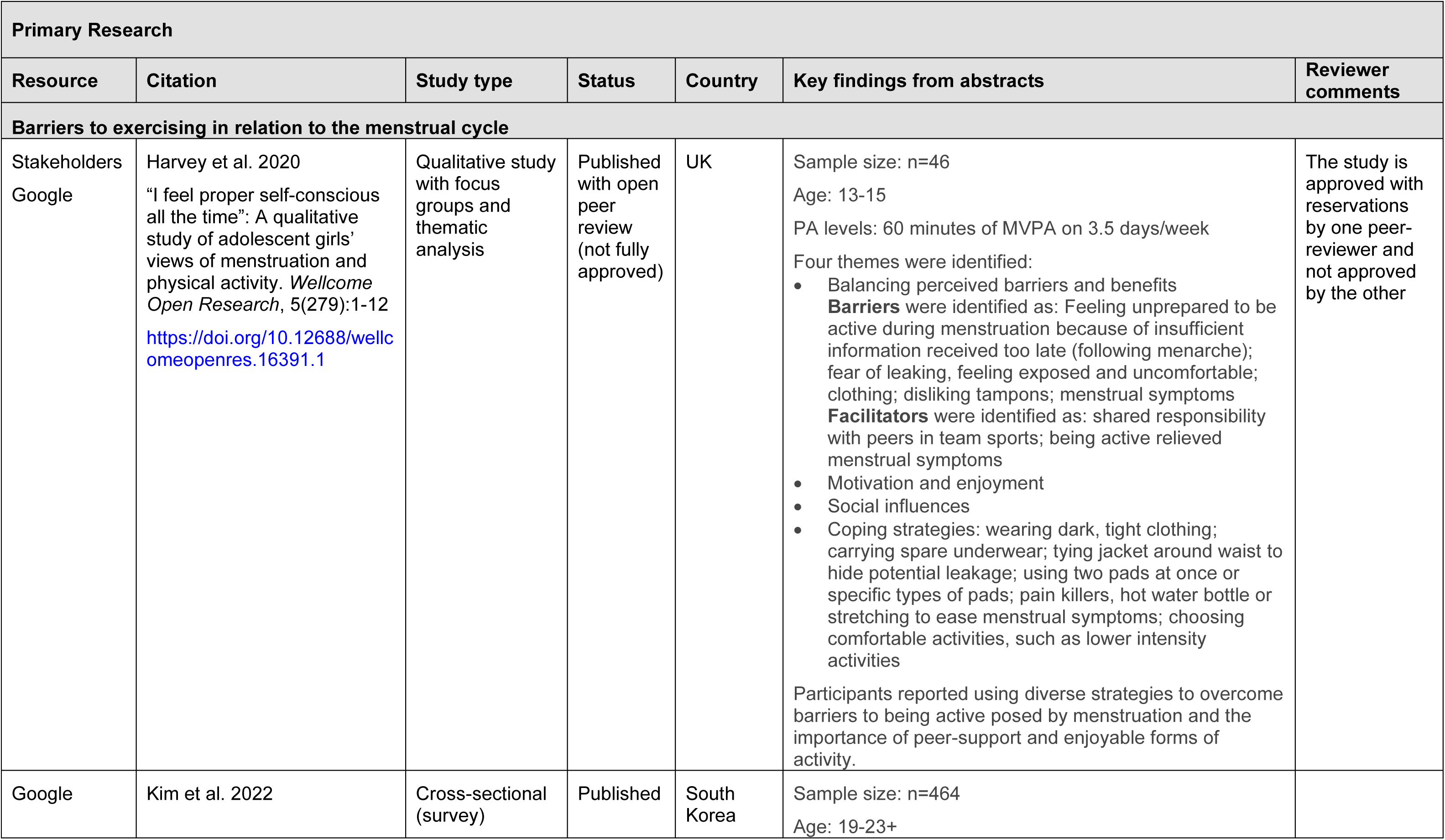
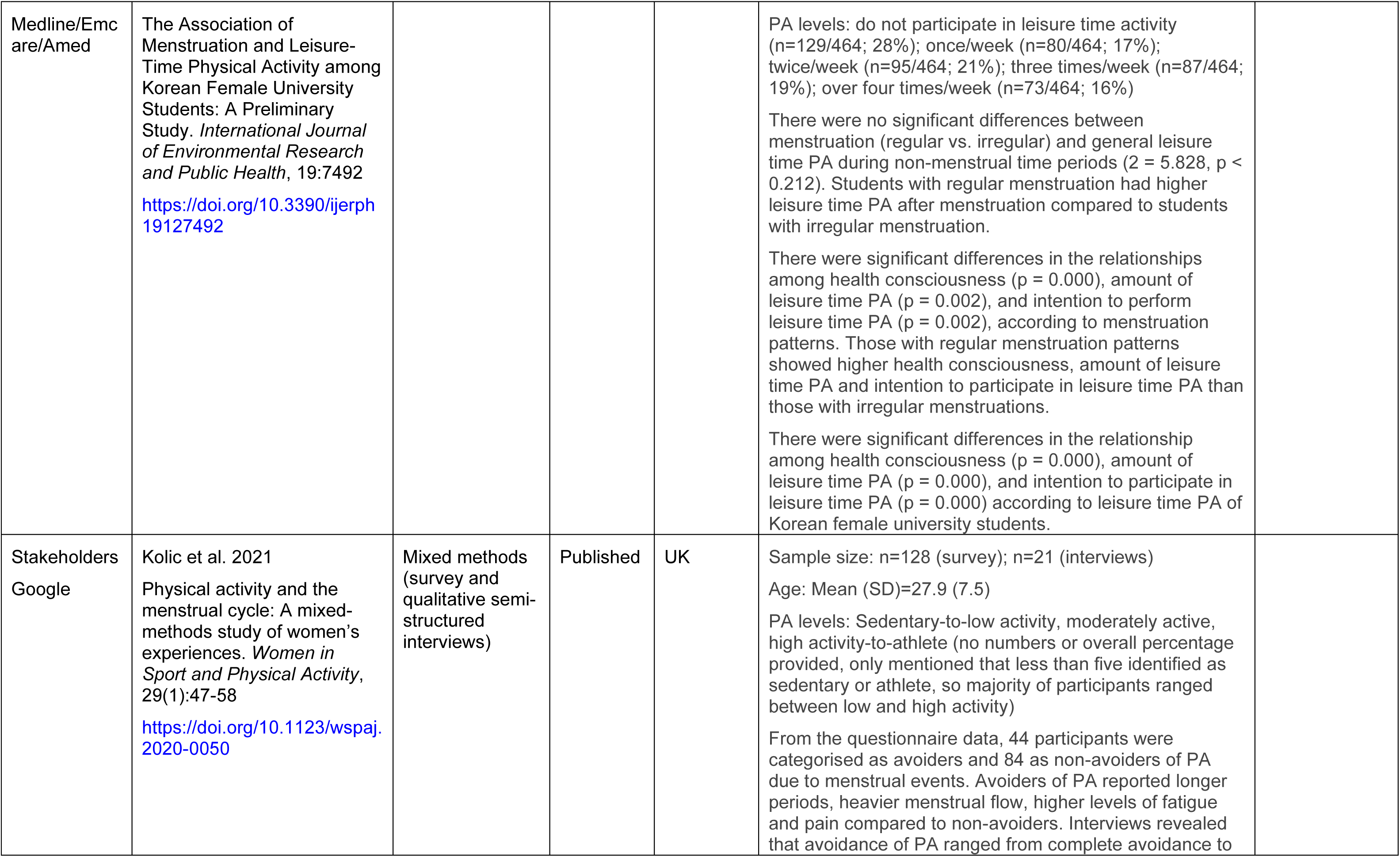
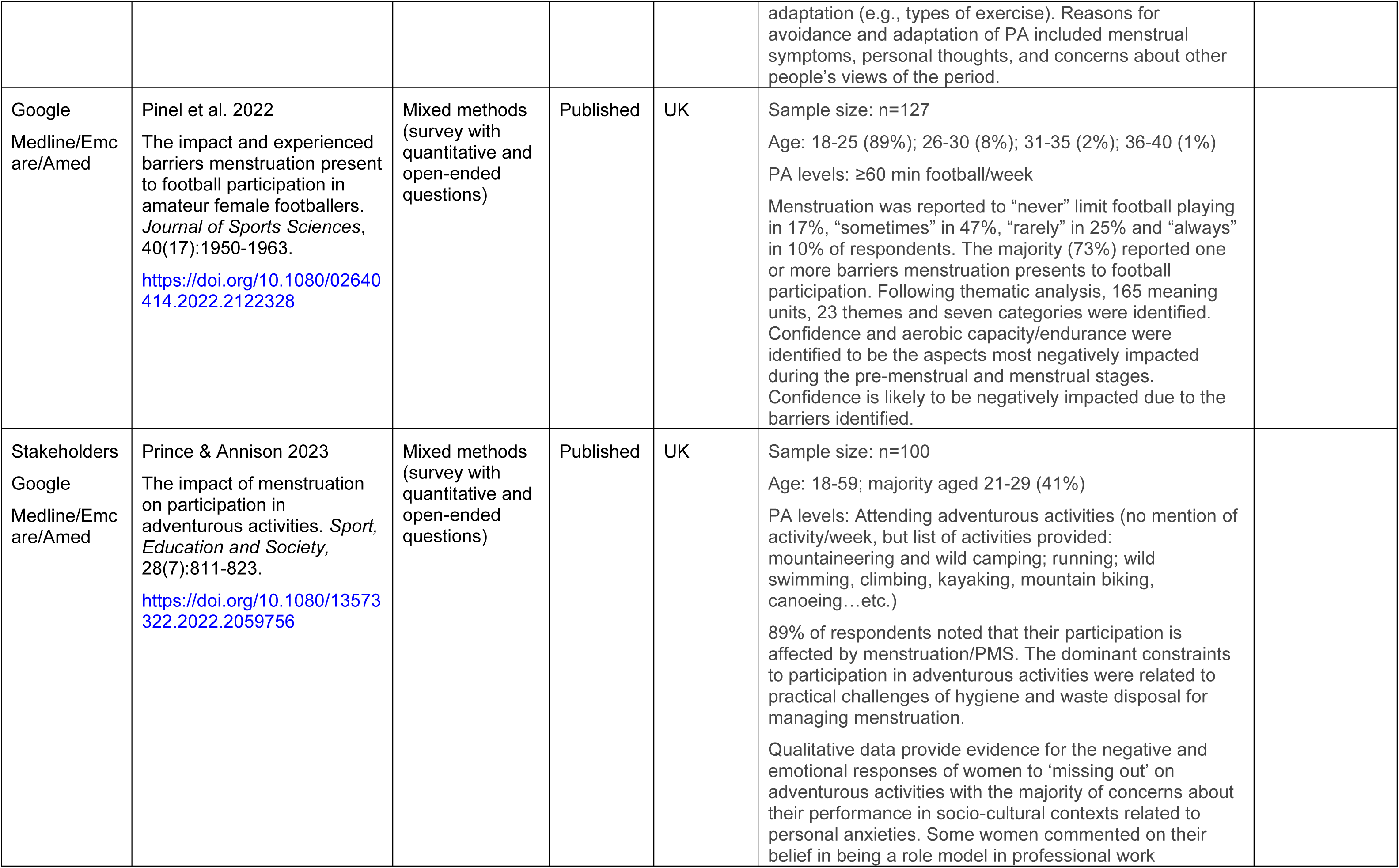
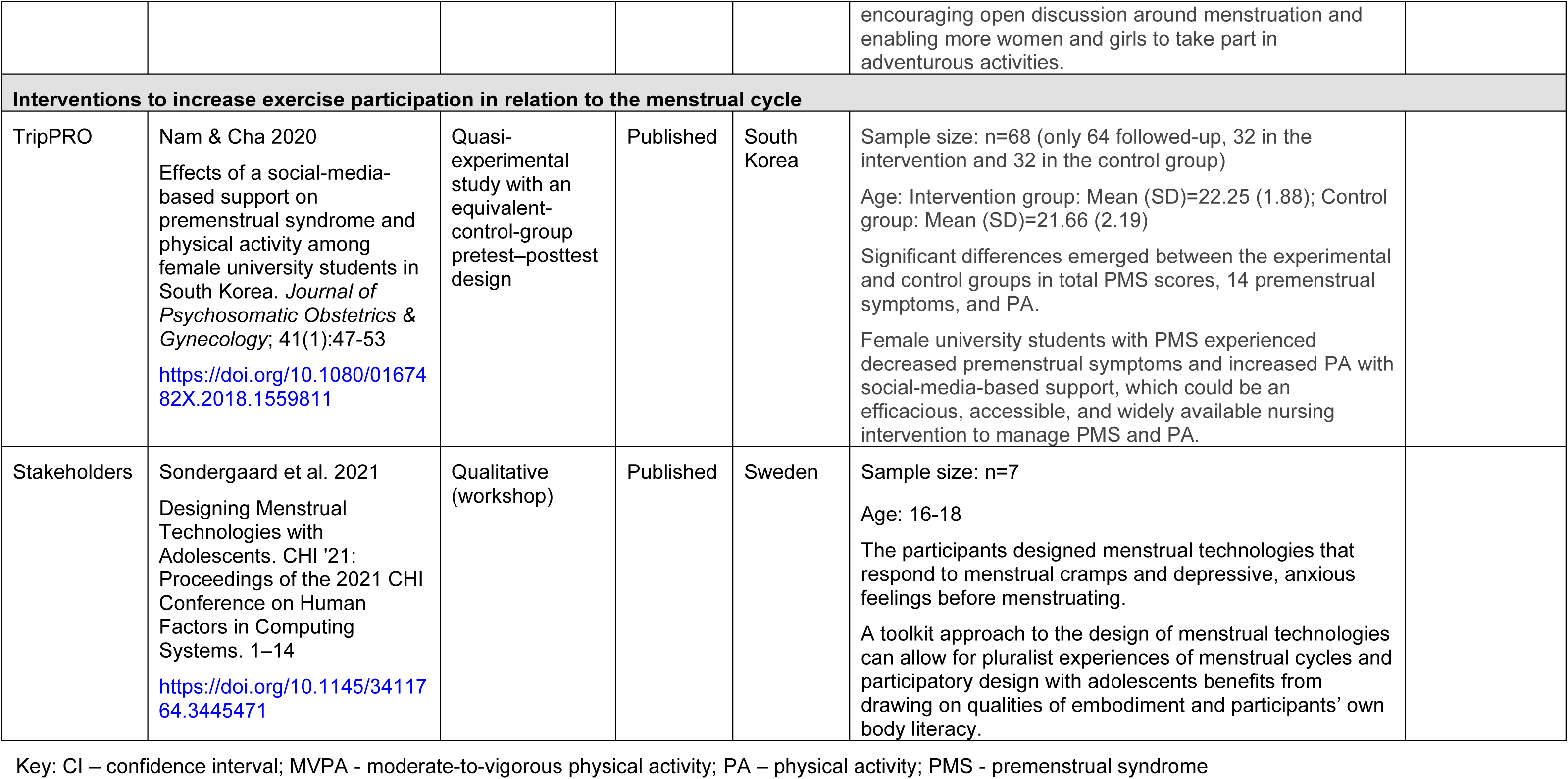
Summary of included primary research evidence.

**Table 4:** Summary of included organisational reports.

| Citation<br>Citation retrieval source | Details |
| --- | --- |
| **Barriers to exercising in relation to the menstrual cycle** |  |
| House of Commons 2024<br>Health barriers for girls and women in sport<br>https://publications.parliament.uk/pa/cm5804/cms/elect/cmwomeq/130/report.html#heading-1<br>Google search | Type of publication: Government report<br>Key findings:<br>*School-aged children*<br>Barriers to girls' participation in sport and exercise begin from an early age and are partly driven by hormonal differences. There is a PE enjoyment gap between girls and boys, which is particularly pronounced at KS4 level, where 59% of girls say they enjoy PE, compared to 84% of boys. Part of the reason for this gap is the differences in how girls and boys experience puberty, with girls facing additional barriers. Thirty-eight percent of secondary school girls surveyed by Youth Sports Trust raised periods as a barrier to participation. Of those who did, 68% were worried about being in pain or discomfort, 60% about leaking, and 57% about low mood while on period.<br>The report noted a lack of education about managing the menstrual cycle and periods, with only 51% of schools in England having the menstrual cycle as part of the curriculum despite it being mandatory, and a lack of education about girls going through puberty among PE teachers. It was also noted that education about the menstrual cycle, periods and PA should happen at a younger age because currently it is delivered too late to be useful: after most girls have already gone through puberty. The report proposed that teachers need to be better trained to deliver education around the menstrual cycle, including how to have effective conversations and to support girls better. In addition, it was recommended that the Department for Education and National Physical Activity Taskforce review guidance for schools on PE kit and ensure that schools permit a wide choice of kit for girls, since expanding kit flexibility and choice is thought to increase comfort and the likelihood of participation.<br>*Women aged 41-60*<br>Perimenopausal and menopausal symptoms are some of the barriers to participation in exercise for women in midlife and beyond. These may occur after experiencing barriers at earlier life stages, such as in early years and during puberty. The report noted that current efforts, such as the Government's "Get Active" strategy, are inadequate for this age group as they lack sufficient analysis of the issues experienced by women in midlife and after, measurable targets, and tailored interventions. |
| Plan International UK 2018<br>Break the Barriers: Girls' experiences of menstruation in the UK | Type of publication: Report<br>Key findings:<br>Plan International UK conducted a literature review, interviews with key informants i.e. politicians, council workers, health professionals, teachers and parents/carers; and focus groups with 84 young people. The focus group participants were 56 females and eight males between the ages of nine and 18, from diverse backgrounds. |
| https://plan-uk.org/file/plan-uk-break-the-barriers-report-032018pdf/download?token=Fs-HYP3v<br>Overton | Girls in the UK had similar experiences to girls globally with universal issues being, stigma, taboo, access to menstrual products and pain management.<br>Most focus groups discussed the impact of menstruation on sport and exercise. The main barrier was the concern of leaking. Some girls thought that girls and others who menstruate should not go swimming while on their period.<br>Other issues highlighted by the focus groups was how periods can impact on participation and attendance at school. Either girls missed school due to effects of menstruation or were not allowed to miss school however ill they felt. The media portrayal of people maintaining high levels of activity whilst menstruating made participants feel bad about themselves and guilt for the need of self-care. |
| The Scottish Parliament: Health, Social Care and Sport Committee 2023<br>Female participation in sport and physical activity<br>https://digitalpublications.parliament.scot/Committees/Report/HSCS/2023/10/9/44f3cbcf-b4f5-429b-822d-93fdd1e60353-1#907b0806-d908-4928-96c4-791b373dcd60.dita<br>Overton | Type of publication: Report<br>Key findings:<br>The report notes that from puberty there is a persistent gender gap in the rates of participation in sport and PA. It discusses the barriers and any progress being made in addressing those barriers.<br>*Barriers faced by teenage girls*<br>Barriers faced by teenage girls include puberty, gendered/restricted offerings, a focus on competition rather than having fun, negative attitudes by boys and boys' domination of playground spaces. Regarding the barriers experienced due to puberty, the Health, Social Care and Sport Committee acknowledges the implementation of the Scottish Government's Women's Health Plan that encourages education about menstrual health as part of the Scottish curriculum. It is hoped that this will improve awareness and understanding of the impact that menstruation can have on girl's participation in PA and remove the stigma around managing periods. The Committee requests an evaluation of the plan's impact.<br>The report also notes that one barrier is being allowed or not to use toilet facilities, and the appropriateness of toilet and changing facilities when a girl is menstruating. Additionally sport kits/uniforms can be a barrier to participation in sport and PA such as white or form-fitting clothing. Whilst the Scottish Government is committed to bring forward new statutory guidance on school uniforms (including for sports and PE) that will increase the use of generic and unisex items of clothing, the Committee calls on the Scottish Government to provide further details of the expected timing for introduction of this new guidance. Moreover, the Scottish Government needs to set out how it will ensure that any new uniform requirements do not exacerbate existing concerns about body image or period leakage.<br>*Barriers faced by women of all ages*<br>The barriers to participation in terms of menstruation were identified as period pain, leakage, and a lack of adequate toilet facilities in sports environments, as well as reproductive health conditions such as endometriosis. During the Committee meeting on 23 May 2023, Baz Moffat from The Well HQ pointed out that, as well as those participating in |
|  | sport and PA directly, challenges with managing periods could equally impact female coaches, umpires, volunteers, and others, acting as a barrier to their participation too. |
| Women in Sport 2021<br>Inspiring women to be active during midlife and menopause<br>https://womeninsport.org/resource/inspiring-women-to-be-active-during-midlife-and-menopause/<br>Backchaining | Type of publication: Report<br>Key findings:<br>A third of women aged 41-60 do not meet the guideline of 150 minutes of exercise per week. A fifth are not achieving 30 minutes per week. In lower socioeconomic groups, 50% of women are meeting guidelines for activity, compared to 72% in the highest groups. Often there are few activities directed towards women in midlife, with little support or encouragement to take up activity.<br>Many women in this age group are coping with perimenopause symptoms, which may include erratic and/or heavy periods, difficulty sleeping, and low mood, resulting in having little energy. Based on a survey, 32% of women going through menopause had a physical or mental health condition lasting 12 months or more. Women also reported experiencing shame and embarrassment around menopause symptoms.<br>Thirty percent of women said they were less active since menopause. At the same time, 71% of women experiencing menopause wanted to be more active and 90% would consider exercise if recommended by a GP or health professional.<br>Among the barriers to being active, the report named limited perception of what PA or exercise can be, self-identity as not sporty, underlying health issues and exhaustion, fear of standing out or being ridiculed, not being able to keep up with everyone else or fear of failure, exercise spaces being “not for them”, shame or embarrassment about weight, worry about doing it alone, fear of judgement, assumption they will not enjoy sport and exercise, not seeing the relevance of exercise to their lives, and the needs of others coming first.<br>The document reported on a four-week exercise trial completed by eight women, during which they spent two weeks considering available activities, completed a PA of their choice in the third week, and repeated it in the fourth week. The findings suggested that women can be helped to overcome barriers to PA by expanding their frame of reference, decreasing practical barriers, increased sense of belonging, creating support networks, and recognition of immediate benefits. The report also recommended engaging employers and health professionals to help women take up PA in midlife. |
| Women in Sport 2022<br>Reframing sport for teenage girls: tackling teenage disengagement<br>https://womeninsport.org/wp-content/uploads/2022/03/2022-Reframing-Sport- | Type of publication: Report<br>Key findings:<br>A survey of over 4,000 adolescent girls and boys aged 11-18 years identified that too many girls are disengaging from sport and exercise in their teens with a large difference between boys and girls. The report notes that 43% of teenage girls say they used to be sporty but no longer are compared to 24% of boys. Barriers include self-belief, capability, body image and periods, particularly for girls who are no longer sporty. Around 7 in 10 avoid being active when on |
| for-Teenage-Girls-Tackling-Teenage-Disengagement.pdf<br>Overton | their period (regularly/sometimes). They avoid exercise because of pain (73%), fear of leakage (62%), tiredness (52%) and self-consciousness (45%).<br>Fathers/father figures are less likely to support their daughters than their sons in being active. The majority of girls, both those who are sporty and those who used to be sporty like competitive sports but lack the opportunity to play sport unless they are really good.<br>The report outlines different ways to engage and support girls depending on whether they were never sporty, used to be sporty or are sporty and outlines 8 principles to success. 1) no judgment, 2) invoke excitement, 3) clear emotional reward, 4) open eyes to what's there, 5) build in to existing habits, 6) give girls a voice and choice, 7) champion what's in it for them, 8) expand image of what sporty looks like. |
| Youth Sport Trust 2023<br>Girls Active - National report: Girls' report<br>https://www.youthsporttrust.org/research-listings/research/girls-active-national-reports<br>Backchaining | Type of publication: Report<br>Key findings<br>Of the 18,516 girls surveyed, 74% enjoyed taking part in PA and 64% enjoyed taking part in PE. This compares with 85% of boys enjoying taking part in PA and 86% enjoying taking part in PE. In secondary school (13,859 girls surveyed), the figures for girls dropped to 71% enjoying taking part in PA and 59% enjoying taking part in PE.<br>Among the barriers to PA at school, 38% of secondary-school-age girls listed being on their period. This option was only available to girls in year 7 and above and in that age group, it was the most commonly listed barrier. As for barriers to PA outside of school, being on their period was mentioned by 21% of girls.<br>Questions about PE and periods were answered by girls in year 7 and above. Of those, 51% always took part in PE while on their period, 23% mostly did, 19% sometimes did, and 7% never did. Only 8% felt very comfortable talking to the PE teacher about periods, 44% felt somewhat comfortable, and 48% were not at all comfortable.<br>When asked about concerns about participating in PE or school sport while on their period, 68% listed being in pain or discomfort, 60% leaking, 57% low mood, 52% low energy, 38% low confidence, 31% feeling self-conscious changing in the changing rooms, 31% others knowing they were on their period, 28% feeling self-conscious participating, 28% their skill level being not as good as usual, and 26% not feeling like they could take part to the expected level.<br>When asked what the school could do to help them in PE when they have their period, 40% of girls listed greater understanding and empathy from teachers, 39% greater flexibility to participate to their ability at this time, 36% better options for PE kit to help them feel more comfortable, 32% access to sanitary products in PE, 23% more support to help them cope in PE with how they feel at this time, 19% offering girls-only PE lessons, 11% more support to help them understand about periods and exercise, and 7% information to their parents about PE and periods. |
|  | Regarding PE kit, 56% of girls would like to have more options and 30% wanted extra-curricular kit to be flexible or own choice. |
| **Interventions to increase exercise participation in relation to the menstrual cycle** |  |
| Women in Sport 2023<br>Big Sister: By girls, for girls: big sister impact report<br>https://womeninsport.org/creating-change/piloting-solutions/the-big-sister-project/<br>Overton | Type of publication: Report<br>Key findings:<br>The aim of the Big Sister project was to empower and support teenage girls (9-15) in 11 deprived areas of the UK to enjoy sport, exercise and PA during puberty and their teenage years. This was done through tackling affordability and economic exclusion and removing emotional and practical barriers relating to puberty and periods to allow girls to participate in sport and exercise. To tackle barriers related to periods, period poverty, and stigma, the 'Hey Girls' project provides free sanitary products for girls in areas of deprivation. 'Hey Girls' are the award-winning 'buy one, donate one' social enterprise. Products and information about periods and the different types of products available were distributed in women's toilets/changing rooms at leisure centres, schools, and community projects.<br>The results of the Big Sister project are reported as having positively impacted girls engagement in sport and PA with 44% of girls doing a lot more exercise, 64% enjoying taking part, 61% feeling motivated to take part and 58% more confident to take part in sport and exercise. In turn this has positively impacted girl's health and wellbeing. |

Key: PA – physical activity; PE – physical education; GP – general practitioner

## 8.2 Acknowledgments

The authors would like to thank Steven Macey, Melanie Matthews, Natalie Brown, and Claire James for their contributions during stakeholder meetings in guiding the focus of the review and interpretation of findings.

## 9. APPENDIX

## APPENDIX 1: Resources searched during Rapid Evidence Summary

A single list of resources has been developed for guiding and documenting the sources searched as part of Rapid Evidence Summary. Not all resources will be searched, depending on relevancy. Some sources will be searched as part of the subsequent Rapid Review (or Rapid Evidence Map).

| Secondary research resources | Success or relevancy of the retrieval |
| --- | --- |
| **Medical and health** |  |
| Cochrane Database of Systematic Reviews (CDSR) | Searched, results found |
| JBI (via OVID) | Searched, nothing found |
| NIHR Journals Library | Searched, nothing found |
| Trip | Searched, results found |
| PROSPERO | Searched, results found |
| Medline (via OVID) | Searched, results found |
| Emcare (via OVID) | Searched, results found |
| AMED (via OVID) | Searched, results found |
| **Public health and social sciences** |  |
| Epistemonikos | Searched, results found |
| **Additional resources searched** |  |
| Google Advanced Search | Searched, results found |
| Overton | Searched, results found |

### Literature Search 1

PROSPERO: 12 March 2024

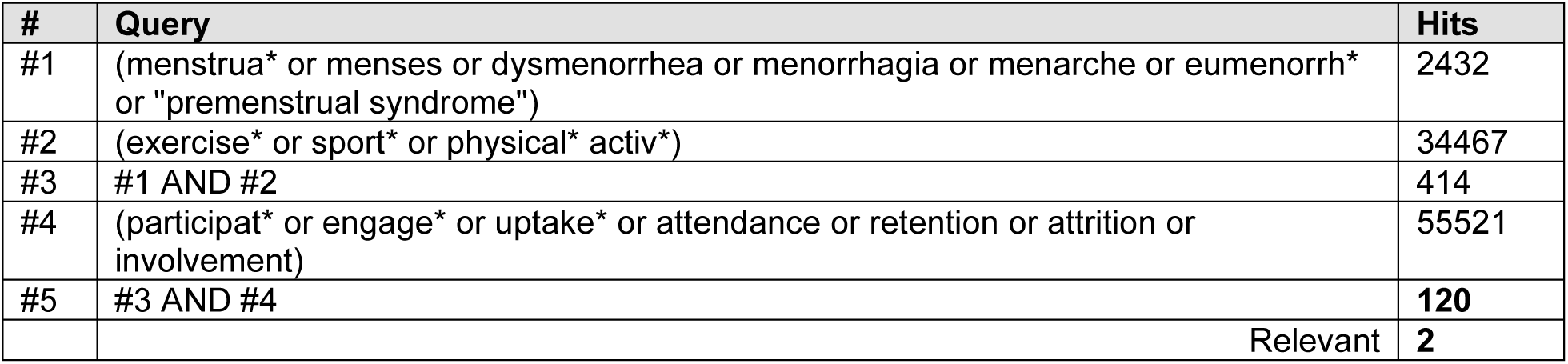

| # | Query | Hits |
| --- | --- | --- |
| #1 | (menstrua* or menses or dysmenorrhea or menorrhagia or menarche or eumenorrh* or "premenstrual syndrome") | 2432 |
| #2 | (exercise* or sport* or physical* activ*) | 34467 |
| #3 | #1 AND #2 | 414 |
| #4 | (participat* or engage* or uptake* or attendance or retention or attrition or involvement) | 55521 |
| #5 | #3 AND #4 | 120 |
|  | Relevant | 2 |

Cochrane Library: 12 March 2024

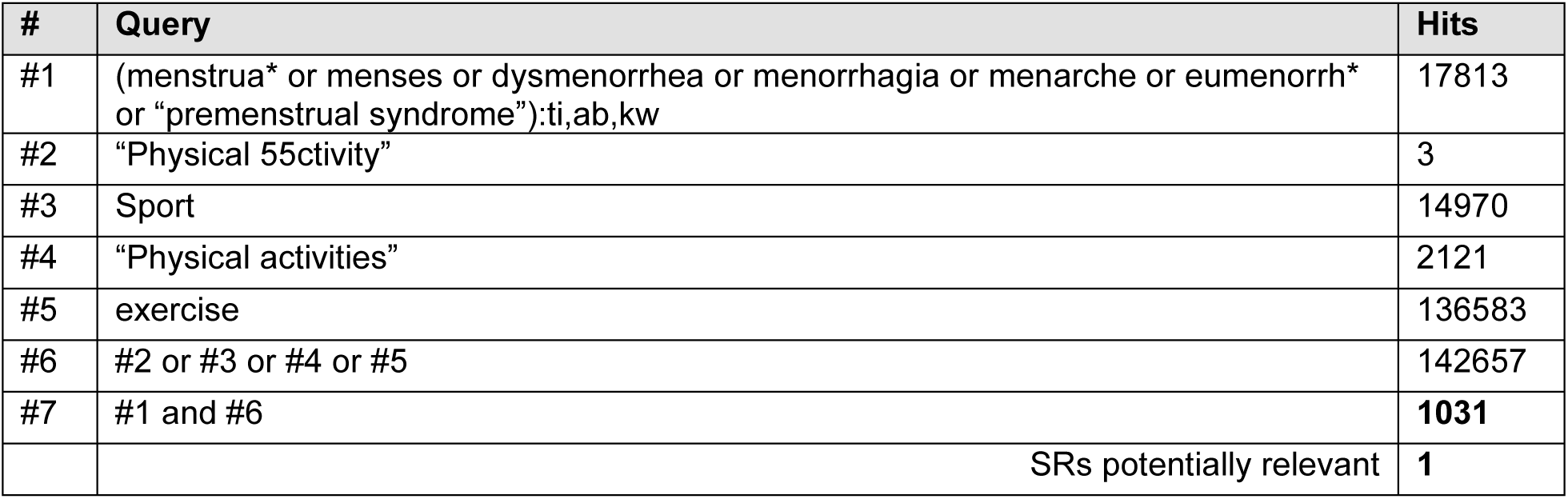

JBI EBP Database: 12 March 2024

JB I EBP Database: 12 March 2024
| # | Query | Hits |
| --- | --- | --- |
| 1 | (menstrua* or menses or dysmenorrhea or menorrhagia or menarche or eumenorrh* or "premenstrual syndrome").mp | 56 |
| 2 | (exercise* or sport* or physical* activ*).mp | 1292 |
| 3 | 1 and 2 | 16 |
|  | Relevant | 0 |

Epistemonikos: 12 March 2024

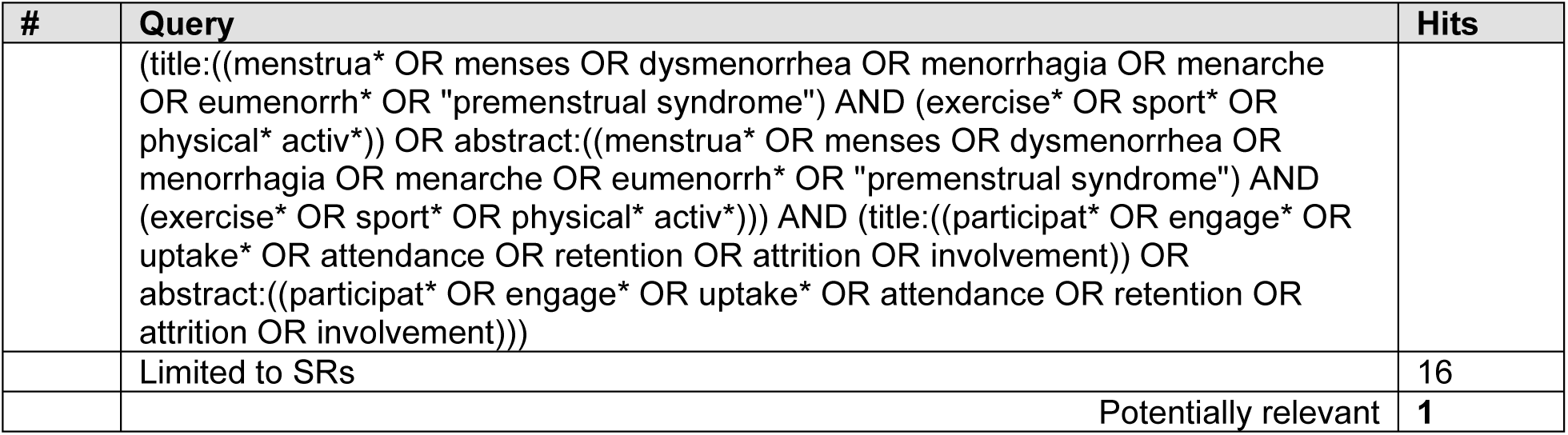

| # | Query | Hits |
| --- | --- | --- |
|  | (title:((menstrua* OR menses OR dysmenorrhea OR menorrhagia OR menarche OR eumenorrh* OR "premenstrual syndrome") AND (exercise* OR sport* OR physical* activ*)) OR abstract:((menstrua* OR menses OR dysmenorrhea OR menorrhagia OR menarche OR eumenorrh* OR "premenstrual syndrome") AND (exercise* OR sport* OR physical* activ*))) AND (title:((participat* OR engage* OR uptake* OR attendance OR retention OR attrition OR involvement)) OR abstract:((participat* OR engage* OR uptake* OR attendance OR retention OR attrition OR involvement))) |  |
|  | Limited to SRs | 16 |
|  | Potentially relevant | 1 |

NIHR Journals Library: 12 March 2024

| Term | Hits |
| --- | --- |
| (menstrua* or menses or dysmenorrhea or menorrhagia or menarche or eumenorrh* or "premenstrual syndrome") AND (exercise* or sport* or physical* activ*) | 27 |
| menstrua* | 29 |
| menses | 0 |
| dysmenorrhea | 0 |
| menorrhagia | 6 |
| menarche | 0 |
| eumenorrh* | 0 |
| "premenstrual syndrome" | 0 |
| Relevant | 0 |

TripPro: 12 March 2024

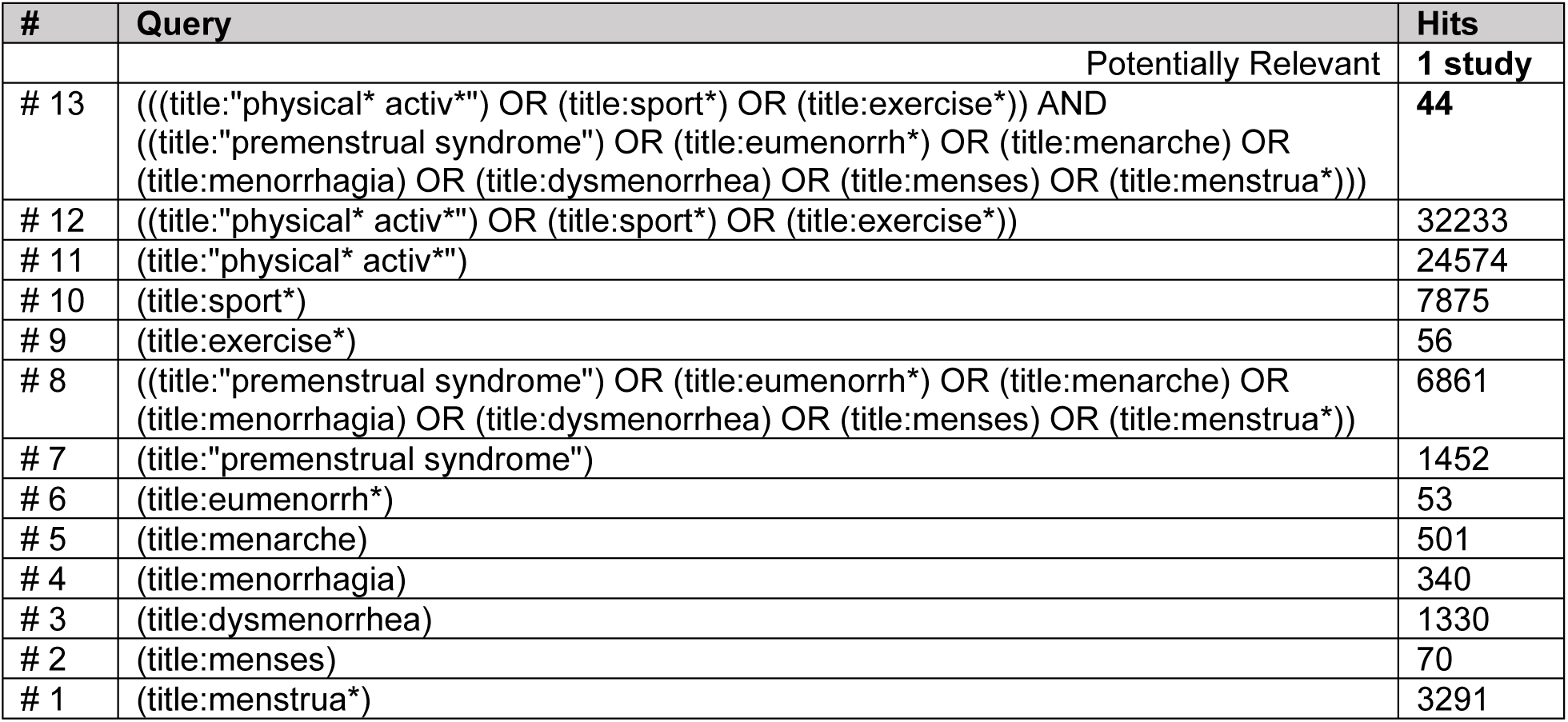

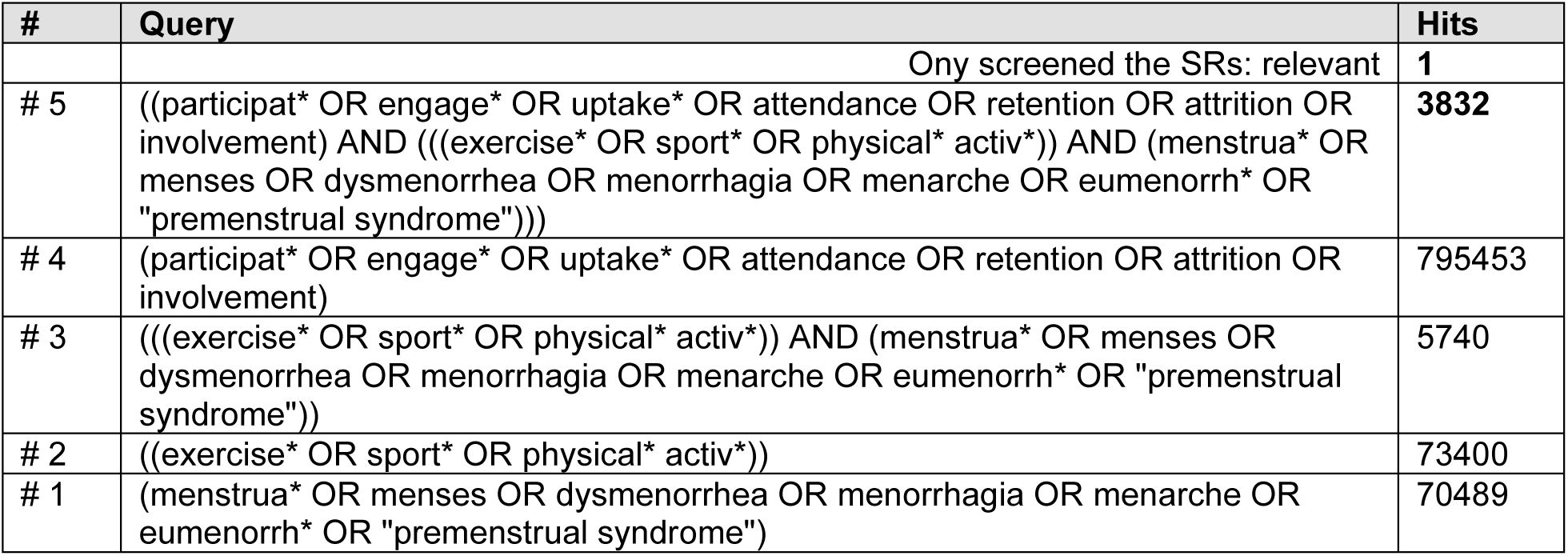

Ovid MEDLINE(R) ALL: 12 March 2024

| # | Query | Hits |
| --- | --- | --- |
| 1 | exp Menstruation Disturbances/ | 29,686 |
| 2 | exp Menstrual Cycle/ | 35,944 |
| 3 | (menstrua* or menses or dysmenorrhea or menorrhagia or menarche or eumenorrh*).tw. | 73,459 |
| 4 | (premenstrual adj (syndrome* or tension*)).tw. | 3,180 |
| 5 | 1 or 2 or 3 or 4 | 103,597 |
| 6 | exp Exercise/ | 254,259 |
| 7 | exp Sports/ | 221,511 |
| 8 | (exercise* or sport* or physical* activ*).tw. | 565,214 |
| 9 | 6 or 7 or 8 | 715,968 |
| 10 | (participat* or engage* or uptake* or attendance or retention or attrition or involvement).tw. | 2,067,777 |
| 11 | **5 and 9 and 10** | **893** |

AMED (Allied and Complementary Medicine): 12 March 2024

| # | Query | Hits |
| --- | --- | --- |
| 1 | exp menstrual cycle/ | 157 |
| 2 | exp Menstruation disorders/ | 663 |
| 3 | (menstrua* or menses or dysmenorrhea or menorrhagia or menarche or eumenorrh*).tw. | 1,019 |
| 4 | (premenstrual adj (syndrome* or tension*)).tw. | 193 |
| 5 | 1 or 2 or 3 or 4 | 1,231 |
| 6 | exp Exercise/ | 10,727 |
| 7 | exp Sports/ | 6,802 |
| 8 | (exercise* or sport* or physical* activ*).tw. | 39,302 |
| 9 | 6 or 7 or 8 | 41,261 |
| 10 | (participat* or engage* or uptake* or attendance or retention or attrition or involvement).tw. | 27,771 |
| 11 | **5 and 9 and 10** | **21** |

Ovid Emcare: 12 March 2024

| # | Query | Hits |
| --- | --- | --- |
| 1 | exp menstrual cycle/ | 15,712 |
| 2 | exp menstruation disorder/ | 13,797 |
| 3 | (menstrua* or menses or dysmenorrhea or menorrhagia or menarche or eumenorrh*).tw. | 21,790 |
| 4 | (premenstrual adj (syndrome* or tension*)).tw. | 1,074 |
| 5 | 1 or 2 or 3 or 4 | 36,901 |
| 6 | exp exercise/ | 143,420 |
| 7 | exp sport/ | 93,719 |
| 8 | exp physical activity/ | 193,088 |
| 9 | (exercise* or sport* or physical* activ*).tw. | 309,629 |
| 10 | 6 or 7 or 8 or 9 | 456,692 |
| 11 | (participat* or engage* or uptake* or attendance or retention or attrition or involvement).tw. | 719,693 |
| 12 | **5 and 10 and 11** | **547** |

| **Database** | **Results** |
| --- | --- |
| Medline | 893 |
| AMED | 21 |
| Ovid Emcare | 547 |
| **TOTAL** | **1461** |
| Duplicates in Endnote | 378 |
| Duplicates in Raayan | 17 |
| **New TOTAL** | **1066** |

### Literature Search 2

PROSPERO: 18 March 2024

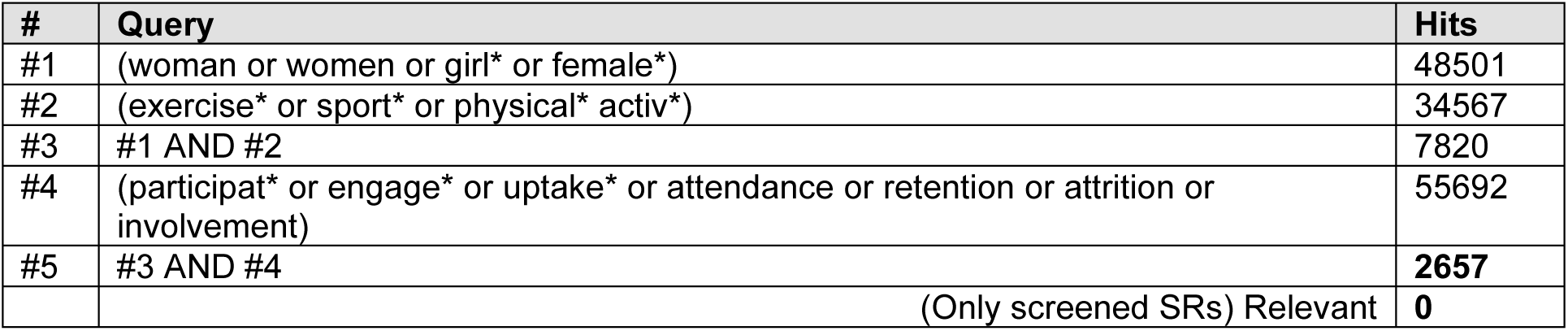

| # | Query | Hits |
| --- | --- | --- |
| #1 | (woman or women or girl* or female*) | 48501 |
| #2 | (exercise* or sport* or physical* activ*) | 34567 |
| #3 | #1 AND #2 | 7820 |
| #4 | (participat* or engage* or uptake* or attendance or retention or attrition or involvement) | 55692 |
| #5 | #3 AND #4 | **2657** |
|  | (Only screened SRs) Relevant | **0** |

Cochrane Library: 18 March 2024

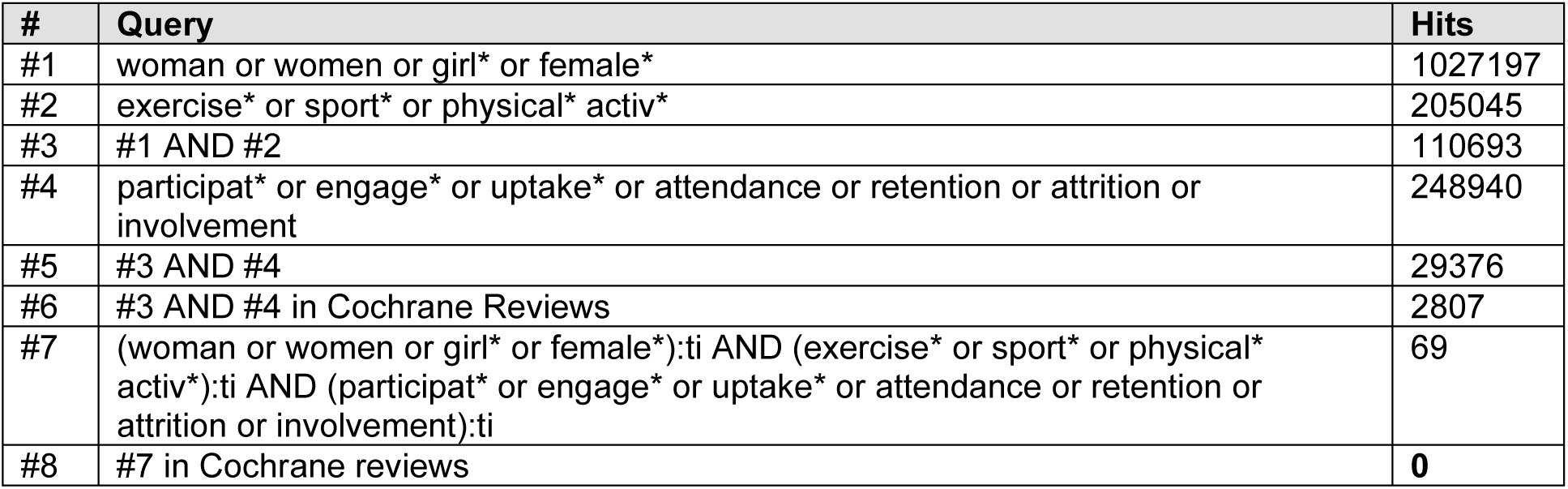

JBI EBP Database: 18 March 2024

| # | Query | Hits |
| --- | --- | --- |
| 1 | (woman or women or girl* or female*).mp. [mp=text, heading word, subject area node word, title] | 1267 |
| 2 | (exercise* or sport* or physical* activ*).mp. [mp=text, heading word, subject area node word, title] | 1292 |
| 3 | 1 and 2 | 325 |
| 4 | (participat* or engage* or uptake* or attendance or retention or attrition or involvement).mp. [mp=text, heading word, subject area node word, title] | 1455 |
| 5 | 3 and 4 | **160** |
|  | Relevant | **0** |

Ovid MEDLINE(R) ALL: 18 March 2024

| # | Query | Hits |
| --- | --- | --- |
| 1 | (wom#n or female* or girl*).ti. | 484,004 |
| 2 | exp Exercise/ | 254,462 |
| 3 | exp Sports/ | 221,655 |
| 4 | (exercise* or sport* or physical* activ*).tw. | 565,709 |
| 5 | 2 or 3 or 4 | 716,562 |
| 6 | (participat* or engage* or uptake* or attendance or involvement).tw. | 1,859,856 |
| 7 | exp "Systematic Review"/ | 255,293 |
| 8 | exp Meta-Analysis/ | 196,945 |
| 9 | exp Systematic Reviews as Topic/ | 12,892 |
| 10 | exp Meta-Analysis as Topic/ | 29,305 |
| 11 | (systematic review* or meta-analysis or review*).pt. | 3,474,729 |
| 12 | (systematic adj3 (review* or overview*)).ti,ab,kf. | 341,238 |
| 13 | (quantitative adj3 (review* or overview* or synthes*)).ti,ab,kf. | 9,059 |
| 14 | (meta-analy* or metaanaly* or meta-synthes#s or metasyntes#s).tw. | 300,484 |
| 15 | rapid review*.ti,ab,kf. | 2,255 |
| 16 | 7 or 8 or 9 or 10 or 11 or 12 or 13 or 14 or 15 | 3,580,898 |
| 17 | **1 and 5 and 6 and 16** | **329** |

AMED (Allied and Complementary Medicine): 18 March 2024

| # | Query | Hits |
| --- | --- | --- |
| 1 | (wom#n or female* or girl*).ti. | 6,378 |
| 2 | exp Exercise/ | 10,727 |
| 3 | exp Sports/ | 6,802 |
| 4 | (exercise* or sport* or physical* activ*).tw. | 39,302 |
| 5 | 2 or 3 or 4 | 41,261 |
| 6 | (participat* or engage* or uptake* or attendance or involvement).tw. | 26,391 |
| 7 | exp Meta analysis/ | 427 |
| 8 | (systematic review* or meta-analysis or review*).pt. | 14,041 |
| 9 | (systematic adj3 (review* or overview*)).ti,ab. | 7,324 |
| 10 | (quantitative adj3 (review* or overview* or synthes*)).ti,ab. | 210 |
| 11 | (meta-analy* or metaanaly* or meta-synthes#s or metasyntes#s).tw. | 3,925 |
| 12 | rapid review*.ti,ab. | 40 |
| 13 | 7 or 8 or 9 or 10 or 11 or 12 | 16,824 |
| 14 | **1 and 5 and 6 and 13** | **10** |

Ovid Emcare: 18 March 2024

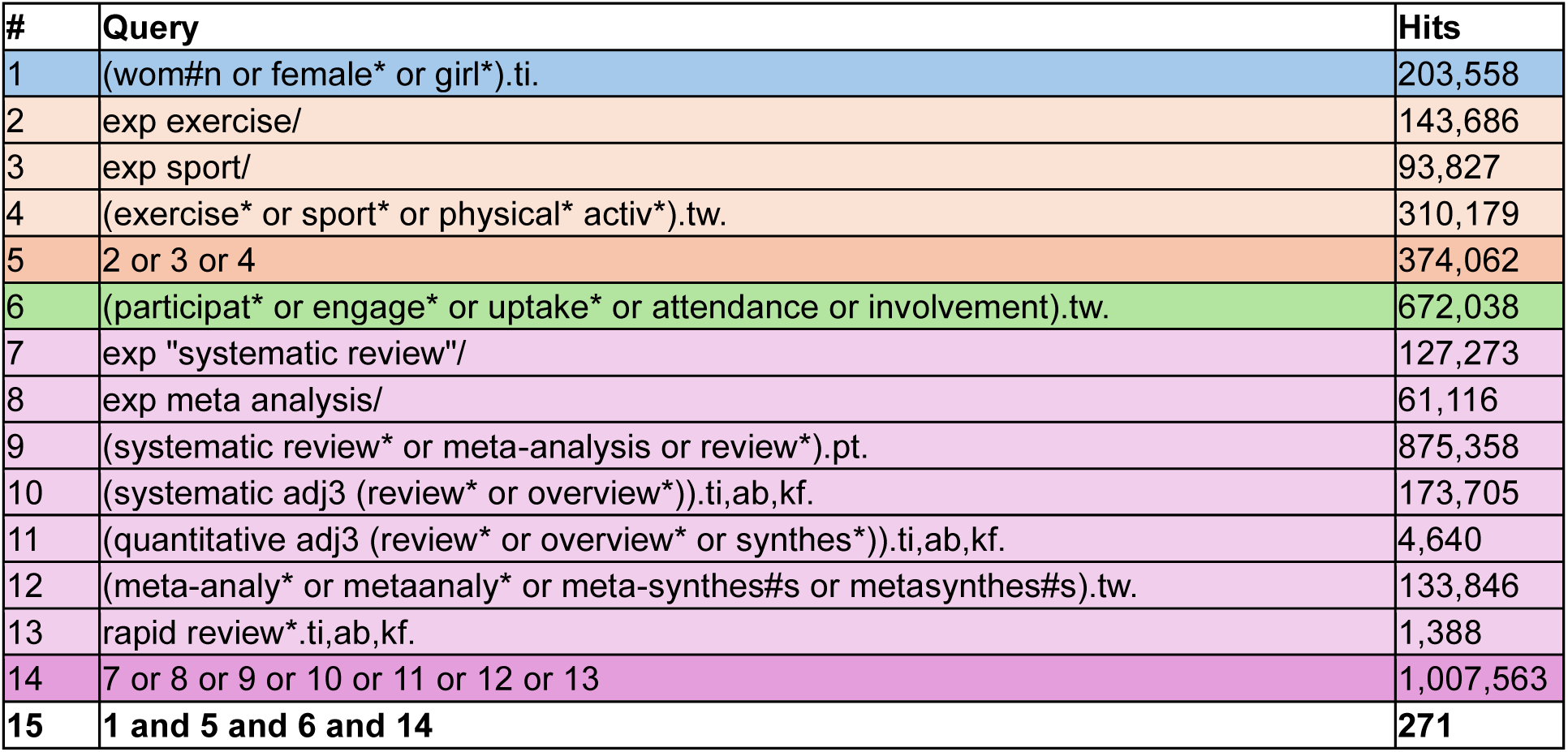

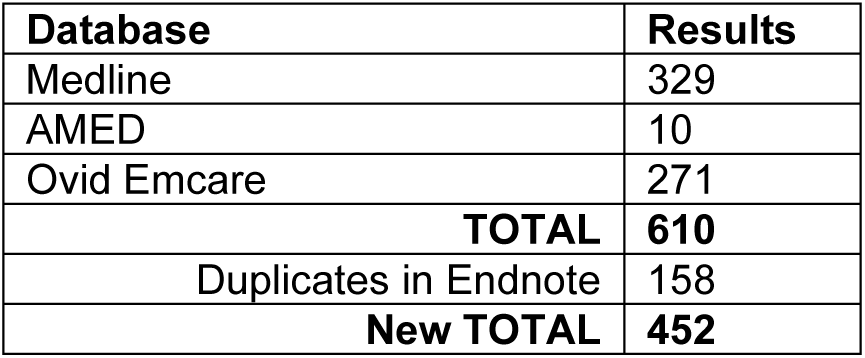

### Additional Google searches for primary research and systematic reviews

| Term | Potentially relevant |
| --- | --- |
| menstruation and barriers to exercise | 5 |
| dysmenorrhea and exercise and systematic review | 1 |
| exercise and barriers and women and systematic reviews | 3 |
| exercise and barriers and menstruation and systematic reviews | 2 |
| aerobic exercise performance and menstruation and systematic review | 5 |
| exercise performance and menstruation and systematic review |  |
| adolescent girls and interventions supporting exercise participation and systematic review | 5 |

